# Identification of Conserved Immune-Related Adverse Event Risk Factors and Clinical Outcomes in a Pan-Immunotherapy Data Mart

**DOI:** 10.1101/2025.06.01.25327978

**Authors:** V.C. Schmid, D.F. Lamparter, R. Mohindra, V. Karanikas, T. Kam-Thong, P. Scepanovic, G. Duchateau-Nguyen, A. Roller, D. Heinzmann, C. Adams, S.L. Mycroft, B.P. Fairfax, N. Städler

## Abstract

**Background:** Cancer immunotherapy often triggers immune-related adverse events (irAEs). Analysis of irAEs in large checkpoint inhibitor (CPI) trials has enhanced their management and demonstrated their prognostic value for treatment outcome. However, data on irAEs in non-standard cancer immunotherapies (CITs) are limited, and systematic exploration is lacking. Identifying predictive biomarkers for irAEs in these therapies is still emerging and essential for improving patient care.

**Methods:** We established a harmonized data mart from 27 early-phase CIT trials, encompassing 14 molecules with diverse mechanisms across various cancer indications. This dataset includes 3,608 patients, both CPI-naïve and CPI-experienced, with detailed information on clinical data, tumor characteristics, soluble biomarkers, and genome-wide genotyping. We examined the occurrence of different irAEs and CIT molecules concerning incidence, severity, and onset. A meta-analysis was conducted to assess the association between risk factors and the time to onset of irAEs. Finally, we explored the predictive value of irAEs for clinical outcomes, specifically measured by progression-free survival (PFS).

**Results:** Our analysis reveals significant variation in irAE incidence and kinetics across CIT molecules. Common irAEs include hepatitis, rash, acute kidney injuries, and hypothyroidism, with hepatitis often severe and others mild. Hepatitis is frequently associated with immunocytokine treatment, while T-cell bispecifics (TCBs) are linked to organ-specific toxicities. Hepatic metastases correlate with hepatitis but inversely with rash; elevated liver enzymes are associated with hepatitis, and high ferritin levels with acute kidney injury risk. Higher myeloid cell counts are associated with reduced rash likelihood. No tumor microenvironment (TME) associations were found, and polygenic risk scores (PGS) show limited utility in our setting. Rash correlates with improved outcomes, whereas hepatitis is associated with a poorer prognosis, independent of baseline prognostic state assessed by the Real World Prognostic score (ROPRO).

**Conclusions:** These findings highlight the complexity of immune toxicities in early-phase trials, emphasizing the importance of the CIT class, as well as the roles of tumor burden, metastasis sites, and systemic immune state in the development of irAEs. Additionally, the observed association between skin toxicities and improved PFS suggests that skin toxicity could serve as a marker of systemic immune activation across immunotherapy contexts.

**Key messages:** What is already known on this topic

● Cancer immunotherapy can induce immune-related adverse events (irAEs); their management and prognostic significance have advanced thanks to data from large checkpoint inhibitor trials.

What this study adds

● This study reveals the complexity of irAEs in early-phase pan-immunotherapy trials, highlighting the impact of tumor burden, metastasis sites, and systemic immune state, while identifying skin toxicity as a potential surrogate marker for improved patient outcomes.

How this study might affect research, practice or policy

● Our study lays a foundation for pan-immunotherapy irAE research, offering insights for clinicians and drug developers to assess risk profiles and guide the design of future trials for new immunotherapies.

## Introduction

Checkpoint inhibitors (CPIs) have significantly enhanced cancer treatment and are now the standard of care for many cancer indications. However, CPI treatment is often associated with the development of immune-related adverse events (irAEs), which vary in type and severity depending on the context and the specific CPI drug administered [1]. Generally, the occurrence patterns of irAEs are known to be highly agent specific. CTLA-4 inhibitors are associated with high-grade irAEs (Grade 3-4) in about 30% of patients, more commonly manifesting as colitis, hypophysitis, and skin rash. In contrast, anti-PD-(L)1 antibody treatment results in fewer severe irAEs, affecting about 10% of patients, with pneumonitis, myalgia, hypothyroidism, arthralgia, and vitiligo being more frequently observed [1,2]. IrAEs typically occur within the first 12 weeks of treatment. They generally occur earlier with anti-CTLA-4 treatment, and distinct timing patterns can be observed across different irAE categories, with cutaneous and gastrointestinal events occurring earlier than endocrine and renal events [2,3]. Additionally, there is evidence suggesting that the occurrence pattern of irAEs depends on the tumor type. It is hypothesized that variations in the tumor microenvironment (TME) may partially explain these tumor-specific differences [1,2]. In a recent study of the PD-L1 inhibitor atezolizumab, the most frequent irAEs of all grades were rash (22.8%), followed by hepatitis (12.4%), hypothyroidism (9.0%), pneumonitis (3.0%), and hyperthyroidism (2.4%). Hepatitis and pneumonitis were the most common high-grade irAEs observed [4]. Although high-grade irAEs are undesirable and may lead to the discontinuation of treatment or, in rare severe cases, death, low grade irAEs are predictive of treatment success [3,5].

Biomarkers that predict the occurrence and severity of irAEs would be invaluable in clinical settings. Despite the extensive data available on CPIs to date, general biomarkers remain elusive and appear to be context-specific [6,7]. Nevertheless, several emerging categories of biomarkers have demonstrated predictive power in various contexts. For example, a larger T-cell receptor repertoire at baseline, along with increased on-treatment clonality and richness, has been associated with the development of irAEs in patients treated with CTLA-4 and PD-1/PD-L1 inhibitors [8–10]. Additionally, elevated levels of effector memory CD4+ T cells at baseline have been observed as predictive markers for irAEs in these treatments. A meta-analysis of 15 Roche-sponsored clinical trials involving atezolizumab assessed baseline risk factors and identified organ-specific risk factors, such as elevated liver enzyme levels for hepatitis and elevated thyroid-stimulating hormone (TSH) for thyroid toxicities. Asian origin was also identified as a risk factor for developing rash, hepatitis, and pneumonitis [4]. Germline genetic biomarkers have been explored as predictors of irAEs. A recent genome-wide association study involving 1,751 patients receiving CPI treatment identified a variant in the *IL7* gene that predisposes individuals to irAEs, which was validated in an independent cohort [11,12]. Similarly, a polygenic risk score (PGS) for hypothyroidism has been associated with the development of hypothyroidism following atezolizumab treatment [13]. The impact of pre-existing auto-antibodies on irAEs has also been investigated. Several studies report an association between increased anti-thyroid antibodies and thyroid dysfunction [14–16]. In conclusion, while general biomarkers for predicting irAEs are still lacking, several promising context-specific biomarkers have been identified for CPI.

In addition to CPIs, T cell engaging bispecific antibodies (TCBs), immunocytokines, and immune modulators have also emerged as promising strategies for the treatment of solid tumors, with several agents rapidly advancing towards clinical application. These innovative CITs are associated with unique adverse event profiles. For instance, two currently approved TCBs for the treatment of solid tumors, Tarlatamab and Tebentafusp, are primarily linked to cytokine release syndrome (CRS) and skin-related events [17,18]. More broadly, TCBs often encounter challenges such as on-target off-tumor toxicity and CRS [19,20]. Furthermore, immunocytokines are another promising class of CITs, with high-dose interleukin-2 (IL-2) being the first FDA-approved treatment for renal cell carcinoma and melanoma. While these first-generation immunocytokines were associated with a substantial side effect burden, the development of second-generation immunocytokines has focused on enhancing their efficacy while reducing or controlling toxicity [21].

Although new patterns of irAEs have been identified in individual clinical trials of non-standard CITs, comprehensive aggregated clinical data remain scarce. The exploration of biomarkers predictive of irAEs is still in its early stages. Therefore, understanding the context and patterns of irAE occurrence, along with identifying biomarkers that can predict these events in novel agents, is crucial for enhancing patient care. To further our understanding of factors influencing irAE risk secondary to non-standard CITs, we have examined both the relationship between tumor characteristics, specific soluble biomarkers, germline genetics and irAEs, as well as the relationship of irAEs with patient outcomes. This analysis is based on harmonized data from 27 Roche-sponsored early-phase clinical trials encompassing 14 molecules in 3,608 patients.

## Methods

### Patient cohort

A harmonized data mart was developed from 27 early-phase CIT trials (26 phase I, 1 phase II) sponsored by F. Hoffmann–La Roche to assess the occurrence of irAEs, identify associated risk factors, and examine their relationship with PFS. This comprehensive dataset includes information from 3,608 patients across trials involving 14 distinct therapeutic molecules. These molecules encompass a range of modalities, such as immunocytokines, immunomodulators, TCBs, costimulatory bispecific antibodies, and agents targeting the PD-1/PD-L1 axis. The trials span various cancer indications, with a primary focus on gastrointestinal cancers (n=725), urinary system cancers (n=504), and lung cancers (n=498). The patient cohort comprises both checkpoint inhibitor (CPI)-naïve and CPI-experienced individuals (n=756), as well as 2,176 patients receiving anti-PD-L1 combination therapies. The dataset offers detailed information on tumor characteristics, including clinical assessments and gene expression profiles, along with data on soluble biomarkers and genome-wide genotyping (see figure 1 and online supplemental table 3).

**Figure 1:**
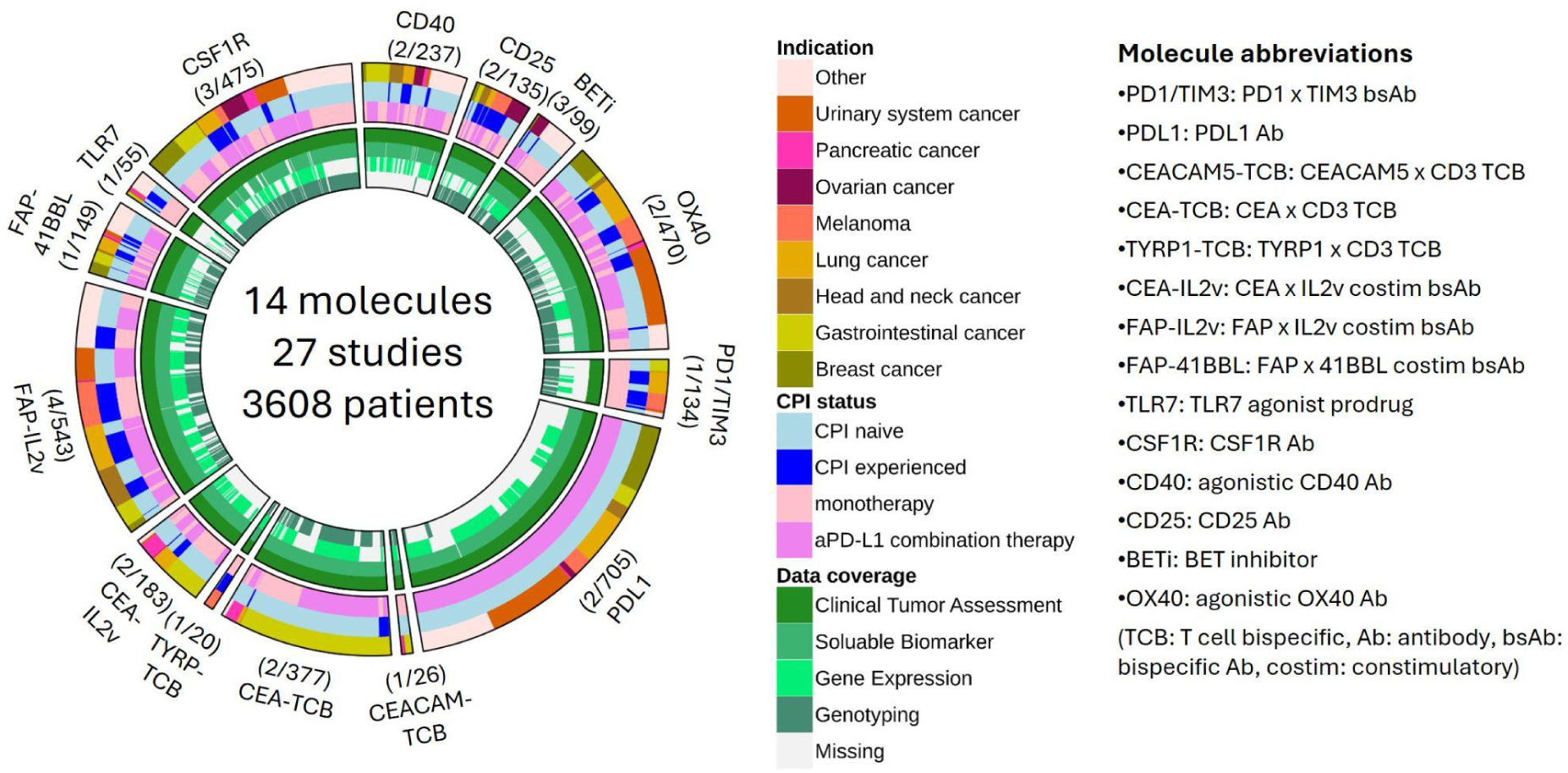
Overview of the harmonized data mart from early-phase CIT trials. The figure illustrates the heterogeneity in CIT treatments and cancer indications, highlighting the depth of data coverage per patient. Note: For each molecule, (X/XX) indicates X number of studies and XX number of patients.

### Immune-related adverse events

irAEs are summarized by medical concept using a comprehensive set of definitions, including Standardized MedDRA Queries, High Level Terms, and Sponsor-defined Adverse Event Grouped Terms [4]. Time to the first adverse event was considered for each irAE category and patient. Safety follow-up times varied across studies (see online supplemental table 4). Competing risks cumulative incidences compared irAE occurrences across studies and molecules, accounting for censoring and varying follow-up times. The NCI CTCAE criteria were used to grade irAE severity. We recorded the most severe event per patient for each irAE category and compared high-grade (grades 3-5) to low-grade (grades 1-2) events both on an individual molecule basis and in aggregate, descriptively without statistical comparisons.

### Risk factors

We evaluated tumor metrics, previous CPI usage, soluble biomarkers, and polygenic risk scores (PGS) as potential risk factors for irAEs. Among the tumor metrics, we assessed liver and lung metastasis status, the number of metastases, the sum of the largest diameter of target lesions (SLD), and tumor microenvironment (TME) features, such as the level of CD8+ T cell infiltration, and immune-related patterns characterized by specific gene signatures. TME features were derived from genome-wide RNA expression data (n=1,900), as previously described [22]. PGS were calculated for patients with available genotyping data (n=1,929) using the EBI Polygenic Risk Score Catalogue [23]. To select PGS for computation, we linked irAE categories to disease-related terms and identified all related subterms using the Experimental Factor Ontology database, which were then mapped to the catalogue. More details on the risk factors, including sample size, distribution, and additional information on genotyping and the polygenic risk score pipeline, are provided in the online supplemental material.

### Statistical analysis

The median follow-up time was estimated using the reverse Kaplan-Meier method [24]. Cumulative incidences of competing risks were calculated, and incidence proportions were compared descriptively without performing statistical comparisons. A two-stage meta-analysis, selected for its robustness and simplicity, was used to identify irAE risk factors. At the study level, Cox regression adjusted for age and sex was applied to assess associations with time-to-event endpoints. A random-effects meta-analysis of log hazard ratios was then used to evaluate the combined effect across all studies. Time-dependent Cox regression was used for time-varying soluble biomarker measurements up to one week from treatment start. Analyses required at least 6 events for Cox regression and 3 log hazard ratios for meta-analysis. Continuous risk factors were log-transformed and standardized at the study level, except for immune-cell proportions, which were logit-transformed. Similarly, two-stage meta-analysis was used to evaluate the association between irAEs and progression-free survival (PFS). Cox regression used PFS as the endpoint and irAE as a time-dependent covariate (’event’,’no event’,’censored’). Log hazard ratios contrasting’event’ and’no event’ were summarized with a random-effects meta-analysis. To adjust the effect of irAEs for the general prognostic state at baseline, a multivariate two-stage meta-analysis was performed, treating PFS as the endpoint and including irAE and the Real World Prognostic score (ROPRO) [25] as covariates in the Cox regression. Finally, to determine the’variance explained’ by polygenic scores (PGS) developed for liver enzymes [26] in relation to corresponding protein measurements, linear regression was performed on baseline enzyme measurements against PGS, stratified by liver metastasis. The regression slopes were combined across studies via meta-analysis and then squared. Only studies with at least 10 samples available in each stratum and at least 40 in total were included. All analyses were performed using R Statistical Software [27].

## Results

### Occurrence of irAEs

Among a total of n=3,573 safety-evaluable patients, n=973 experienced at least one instance of hepatitis, n=967 had rash, n=366 had acute kidney injury, n=159 had hypothyroidism, n=81 had pneumonitis, n=59 had colitis, n=55 had pancreatitis, and n=43 had hyperthyroidism (see table 1). A significant proportion of the hepatitis, pneumonitis, colitis, and pancreatitis cases were severe (grades 3-5), with 45.2%, 39.0%, 48.3%, and 50.9% of cases, respectively. In contrast, rash, acute kidney injury, hypothyroidism, and hyperthyroidism were predominantly mild (grades 1-2), with 7.6%, 14.0%, 2.6%, and 4.9% of cases, respectively (see table 1). The severity of irAEs appears to be influenced mainly by the irAE category and, to a lesser extent, by the CIT treatment (see online supplemental figure 3).

**Table 1:** Summary of irAEs: n represents the number of patients experiencing at least one irAE; N denotes the total number of patients evaluated for safety; n_lowgrade indicates the number of patients with at least one low-grade event (grades 1-2); n_highgrade refers to the number of patients with at least one high-grade event (grades 3-5).

|  | Total events |  |  | High vs low grade events |  |  |
| --- | --- | --- | --- | --- | --- | --- |
|  | n | N | incidence | n_lowgrade | n_highgrade | percentage high grade |
| Hepatitis | 973 | 3573 | 27.2 | 527 | 435 | 45.2 |
| Rash | 967 | 3573 | 27.1 | 865 | 71 | 7.6 |
| Acute Kidney Injury | 366 | 3573 | 10.2 | 313 | 51 | 14 |
| Hypothyroidism | 159 | 3573 | 4.5 | 151 | 4 | 2.6 |
| Pneumonitis | 81 | 3573 | 2.3 | 47 | 30 | 39 |
| Colitis | 59 | 3573 | 1.7 | 30 | 28 | 48.3 |
| Pancreatitis | 55 | 3573 | 1.5 | 26 | 27 | 50.9 |
| Hyperthyroidism | 43 | 3573 | 1.2 | 39 | 2 | 4.9 |

Next, we assessed the incidences of irAEs for different molecules. To account for varying study follow-up times, we examined the incidence proportion over a 6-month period (figure 2A). The incidence proportion substantially varies across different CIT molecules, with the most prevalent irAE being hepatitis (21.4-34.9%), rash (19.2-42.6%), acute kidney injury (7.9-13.5%), and hypothyroidism (0.5-7.9%), all reported as 25th to 75th percentiles. Hepatitis occurs more frequently with systemic immunocytokines CEA-IL2v and FAP-IL2v (46.2% and 60.2%, respectively) and related immune stimulators FAP-41BBL and TLR7 (38.8% and 35.0%, respectively). Targeted treatments with TCBs are linked to organ-specific toxicities, such as rash and colitis for CEA-TCB (42.7% and 4.4%, respectively), rash for TYRP1-TCB (61.4%), and pancreatitis for CEACAM5-TCB (29.2%). CD25 antibody treatment is associated with high incidences of rash (59.5%), while BET inhibitors are linked to higher incidences of acute kidney injury (18.6%).

**Figure 2:**
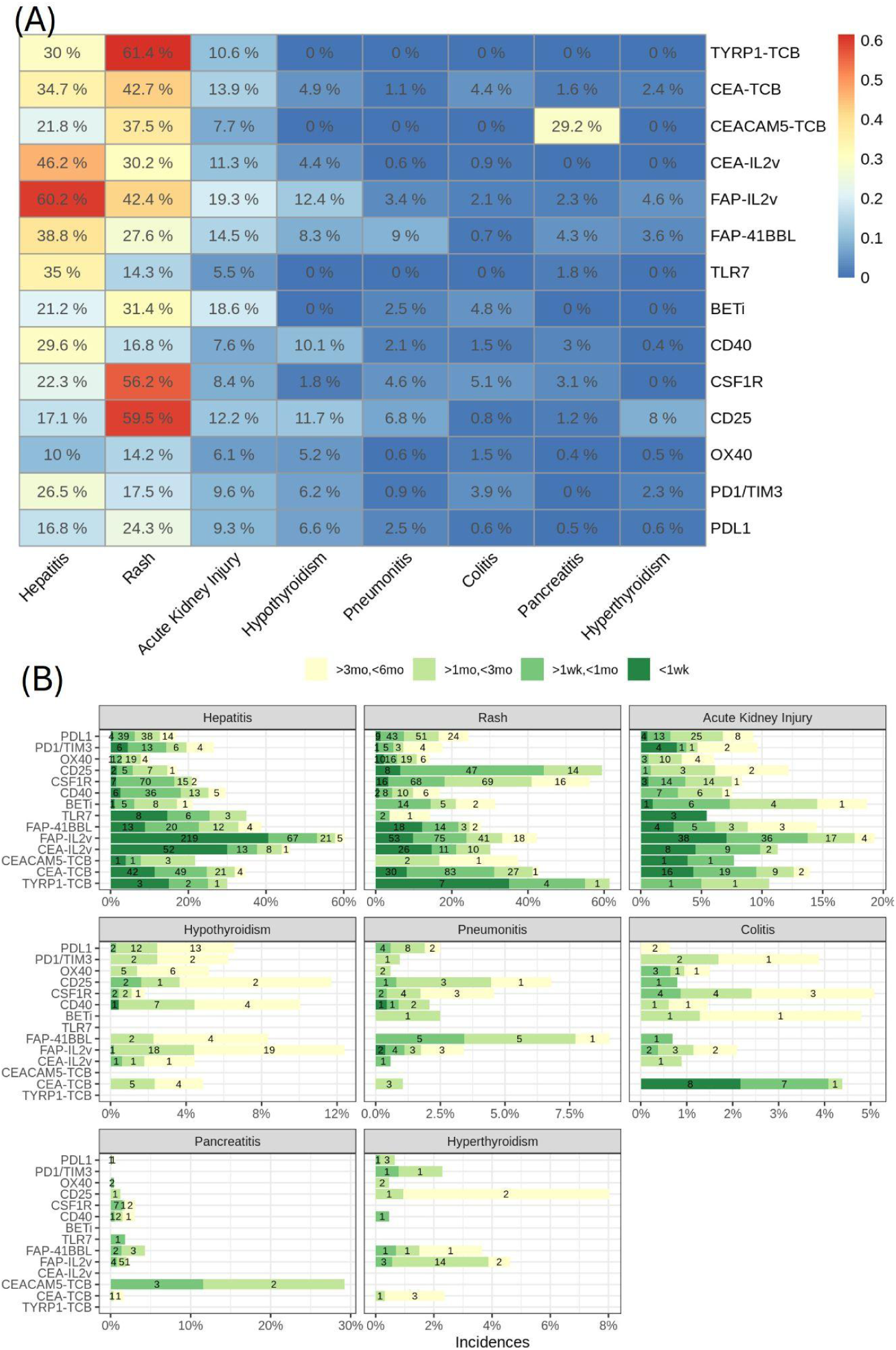
Incidence and Onset of irAEs. (A) Six-month incidence proportions of irAEs for various CIT molecules, with cumulative incidences represented by numbers and colors. (B) Onset timing of irAEs categorized by CIT molecules, showing cumulative incidences up to 6 months: instant (<1 week), rapid (1 week to 1 month), moderate (1 to 3 months), and delayed (3 to 6 months), along with the number of events in each category.

Finally, we assessed the kinetics of different irAEs and CIT molecules by categorizing them into instant (<1 week), rapid (1 week to 1 month), moderate (1 to 3 months), and delayed onset (3 to 6 months). The onset of irAEs varies greatly across treatments and irAE types. Notably, early onset appears to be characteristic of hepatitis, specifically for IL-2 immunocytokines where the onset occurs instantly in the majority of cases, whereas late onset seems to be associated with hypothyroidism (see figure 2B).

In the following sections, we will focus on the most frequent irAEs: hepatitis, rash, acute kidney injury, and hypothyroidism.

### Identification of irAE risk factors

Next, we assessed tumor metrics, previous CPI usage, soluble biomarkers, and polygenic risk scores as potential risk factors for irAEs. Each factor was tested for association with irAEs using a confounder-adjusted two-stage meta-analysis. Specifically, for each irAE category and potential risk factor, the association was assessed at the study level using Cox regression adjusted for age and sex. The obtained hazard ratios were then summarized using a random-effects meta-analysis. The resulting hazard ratios (HR) and 95% confidence intervals (CI) are displayed in figures 3, 4, and 5. We further stratified the two-stage meta-analysis by concomitant anti-PD-L1 use to assess its impact. Results are in the online supplemental material.

**Figure 3:**
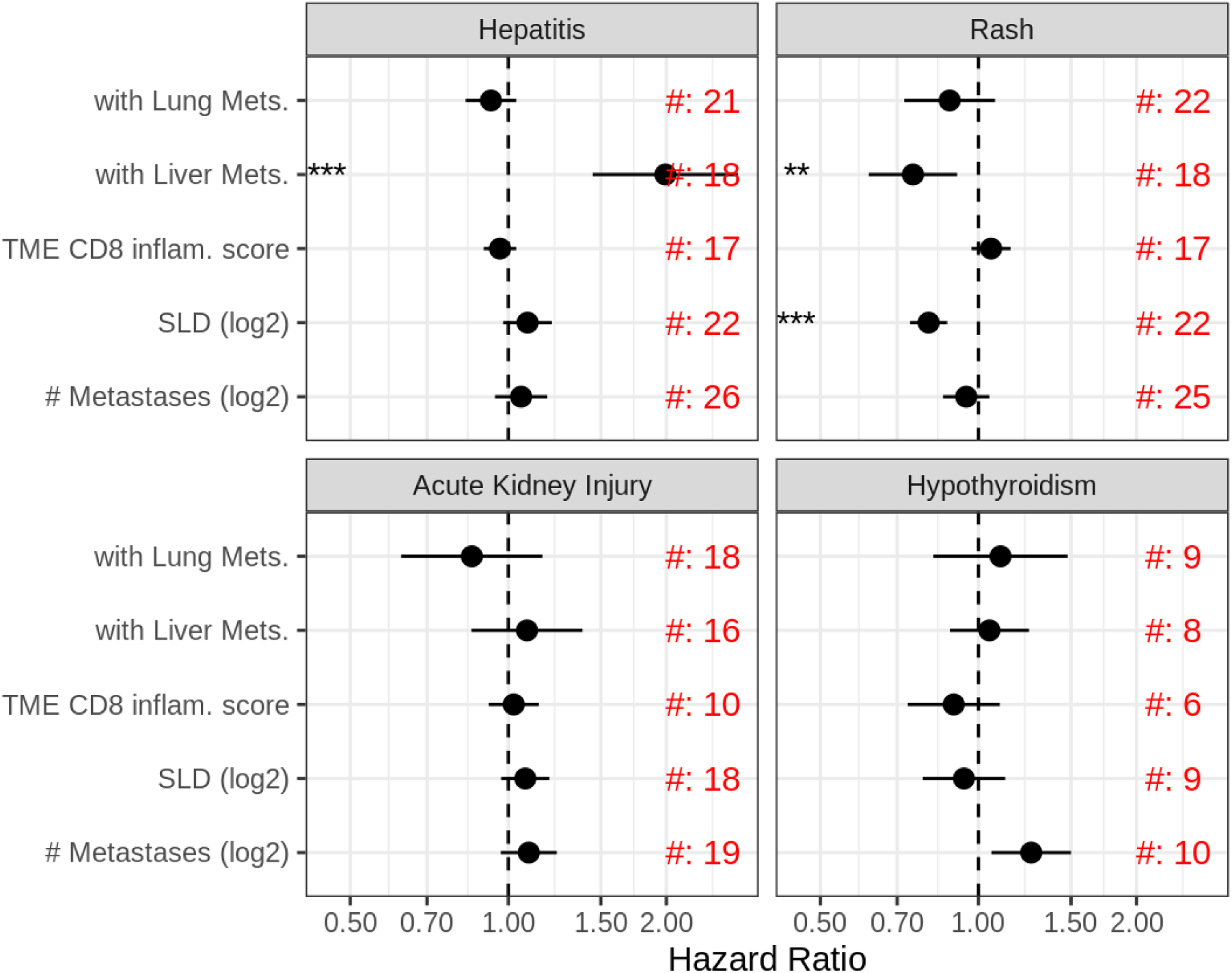
Association of tumor metrics with irAEs. Tumor metrics include the presence of liver and lung metastases (Liver/Lung Mets), the number of metastases (log2-transformed), the sum of the largest diameters of target lesions (SLD, log2-transformed), and the level of CD8+ T cell infiltration (TME CD8 inflammation score as described [22]). Hazard ratios (HR) and 95% confidence intervals (CI) were obtained through a confounder-adjusted two-stage meta-analysis. #[number] indicates the number of studies included in the meta-analysis. ** indicates results with a p-value <0.01, and *** indicates results with a p-value <0.001.

### Tumor metrics

Among the tumor metrics, we assessed liver and lung metastasis status, the number of metastases, the sum of largest diameter of target lesions (SLD), and tumor microenvironment (TME) features, such as the level of CD8+ T cell infiltration and immune-related patterns characterized by specific gene signatures [22]. Meta-analysis reveals that patients with liver metastases have an increased risk of hepatitis (HR: 1.99, CI [1.45, 2.74]). The same group exhibits a reduced risk of rash (HR: 0.75, CI [0.62, 0.91]), as do patients with smaller SLD at baseline (HR: 0.80, CI [0.74, 0.87]) (see figure 3). We found no evidence suggesting that tumor metrics influence other irAE categories, such as acute kidney injury and hypothyroidism. Additionally, after testing 10 gene expression-based TME features, we found none associated with irAE development, suggesting that irAE development is relatively independent of the TME. For detailed gene signature results, please refer to online supplemental figure 4.

### Previous CPI treatment

We assessed whether previous treatment with CPI impacted the risk of developing irAEs. Out of 3,608 patients, 756 were CPI-experienced. The meta-analysis highlighted moderate evidence for a reduced risk of hepatitis (HR: 0.71, CI [0.55, 0.91]) and hypothyroidism (HR: 0.38, CI [0.19, 0.76]) associated with prior CPI exposure (see online supplemental figure 5).

### Soluble biomarkers

For soluble biomarkers, data were available both at baseline and during treatment, although sampling frequency and data coverage varied significantly across studies (see online supplemental figure 15). To distinguish baseline effects from on-treatment effects, we conducted the meta-analysis twice: once using baseline data only and once incorporating on-treatment data as a time-dependent covariate. Our results on baseline data align with expectations and are consistent with previous findings (see figure 4). Elevated levels of liver enzymes are associated with an increased risk of hepatitis diagnosis, as indicated by an HR of 1.56 for aspartate aminotransferase (AST) with a CI of [1.38, 1.76]. Similarly, thyroid-stimulating hormone (TSH) shows an increased risk of hypothyroidism, with an HR of 1.61 and a CI of [1.36, 1.91]. Higher levels of myeloid cells and C-reactive protein (CRP) (e.g., HR for CRP: 0.82, CI [0.73, 0.91]) are associated with a reduced risk of rash, whereas elevated ferritin levels (HR: 1.38, CI [1.19, 1.60]) correspond to a higher risk of acute kidney injury. Including on-treatment data particularly strengthens the observed association for hepatitis, as exemplified by the increase in the HR for AST from 1.56 to 1.73 (see figure 4). Liver enzymes and thyroid-stimulating hormone are used to diagnose hepatitis and thyroid toxicities, respectively, so the consistent associations we observe are expected.

**Figure 4:**
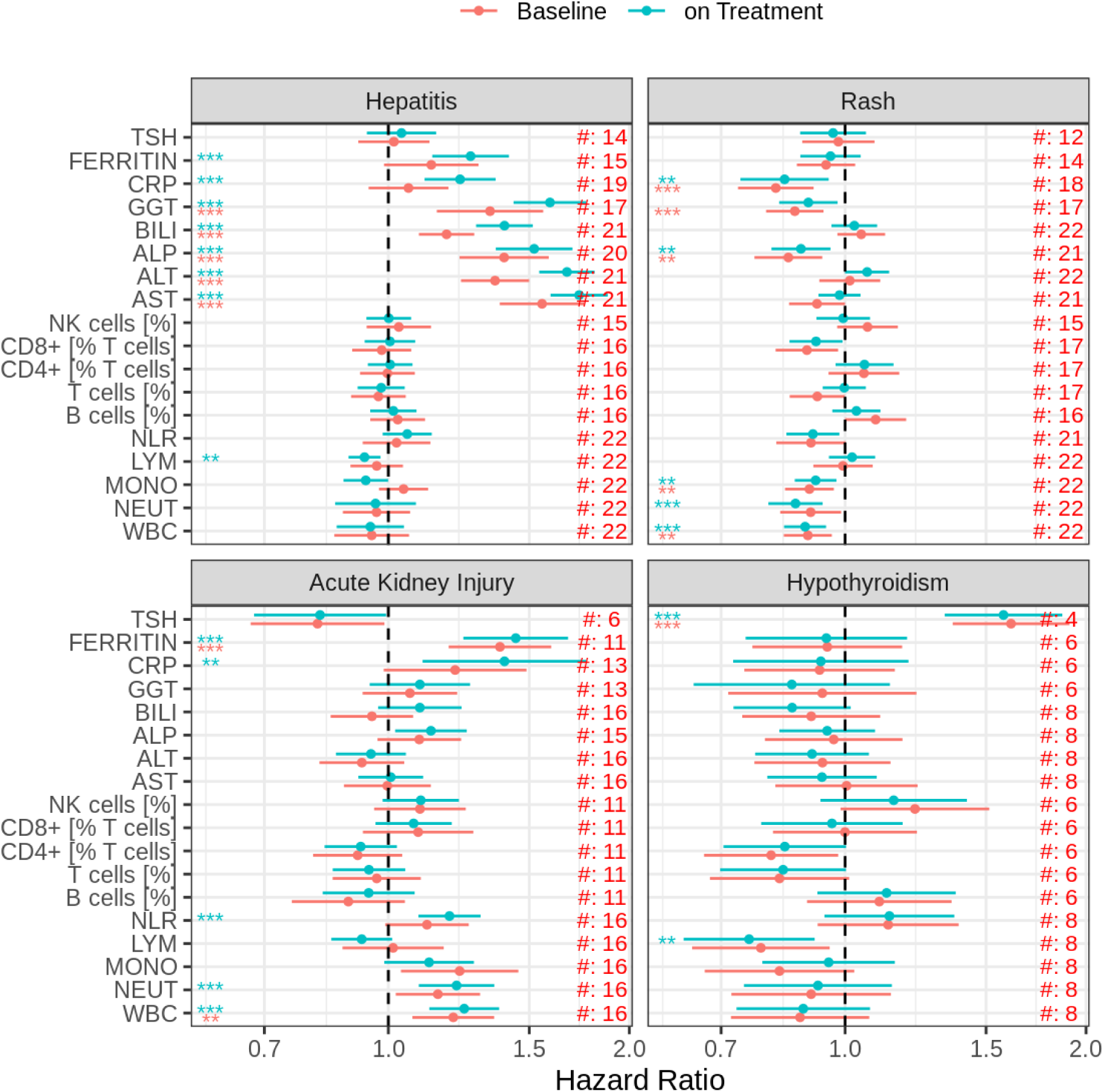
Association of soluble biomarkers with irAEs. Hazard ratios (HR) and 95% confidence intervals (CI) were obtained through a confounder-adjusted two-stage meta-analysis. Red lines represent results obtained from baseline samples only, while blue lines represent results including on-treatment samples. #[number] indicates the number of studies included in the meta-analysis. ** indicates results with a p-value <0.01, and *** indicates results with a p-value <0.001.

### Polygenic risk scores

To assess the presence of germline genetic components predisposing to irAEs, PGS matching irAE categories were calculated for patients with available genotyping data from blood (n=1,929). We tested the association between irAEs and relevant PGS using the aforementioned two-stage meta-analysis approach. The observed associations were generally weak, with only one liver disease score demonstrating a positive association with hepatitis (unadjusted p-value: 0.03; HR: 1.11, CI [1.01, 1.22]). Refer to figure 5A and supplemental table 16 for details. Notably, the PGS developed for liver enzymes—ALP, ALT, AST, and GGT—did not show an association with hepatitis, despite strong signals observed at the protein level. We therefore sought to directly assess the association of PGS with soluble biomarkers and the impact of cancer-induced variability by conducting the analysis for patients with and without liver metastasis. As expected, liver enzymes showed a significant association with their corresponding PGS. Interestingly, the variance explained tended to be larger in patients without liver metastasis, exemplifying the impact of cancer-induced variability. Overall, the maximal observed variance explained was no greater than 4% (see figure 5B). Results for other irAEs are provided in the online supplemental material.

**Figure 5:**
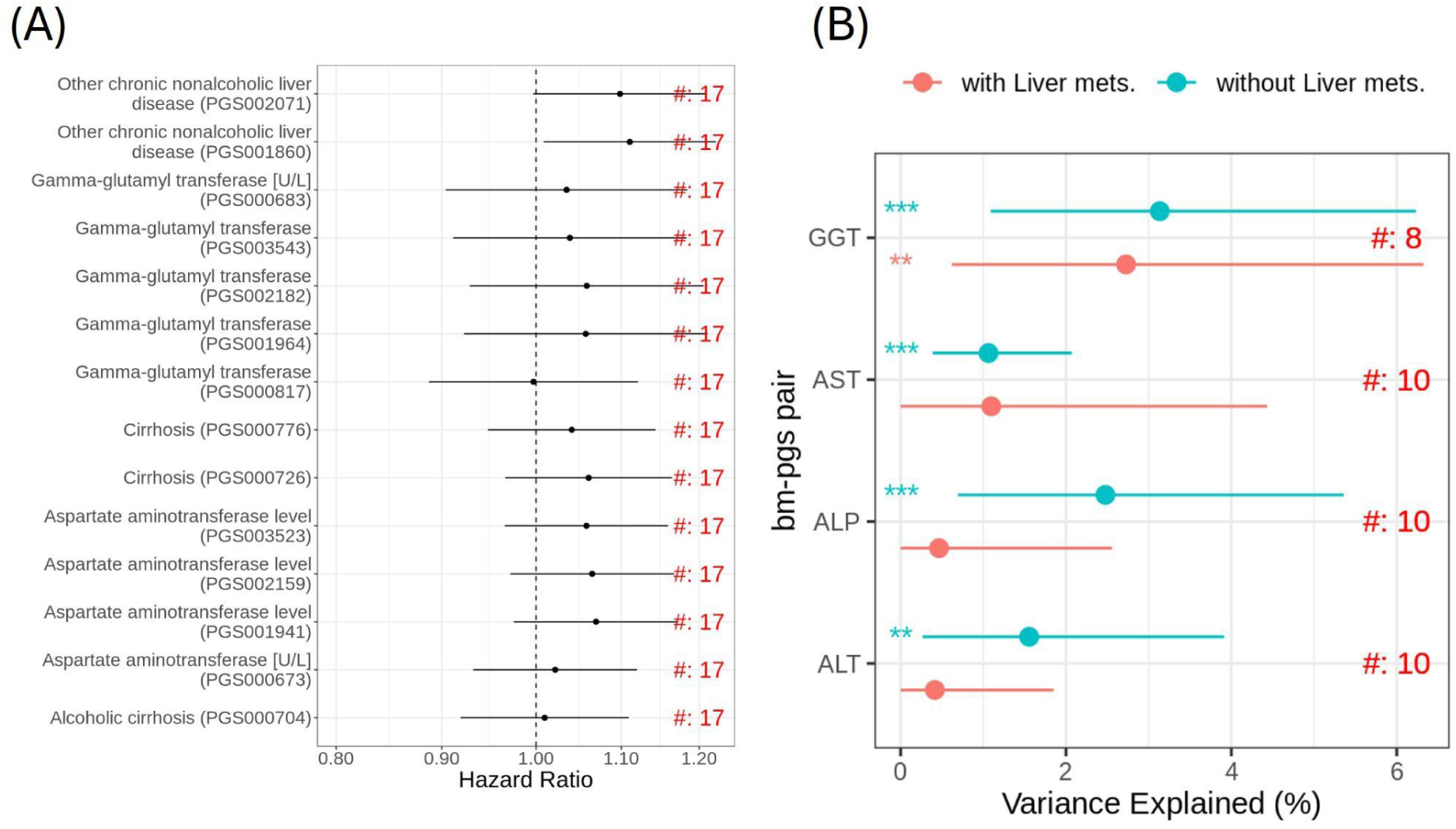
Association with PGS linked to irAEs. (A) Hazard ratios (HR) and 95% confidence intervals (CI) obtained through a confounder-adjusted two-stage meta-analysis for PGS linked to hepatitis. (B) Variance explained by PGS developed for liver enzymes with corresponding protein measurements, stratified by liver metastasis status. Variance explained and 95% CI were obtained through univariate two-stage meta-analysis. #[number] indicates the number of studies included in the meta-analysis. ** indicates results with a p-value <0.01, and *** indicates results with a p-value <0.001.

### Relationship of irAEs with clinical outcome

To assess the relationship between irAEs and PFS, a two-stage meta-analysis was employed. Specifically, Cox regression with PFS as the time-to-event endpoint and irAE as a time-dependent covariate was used to evaluate the association at the study level. The resulting hazard ratios were then summarized using random-effects meta-analysis and are displayed in figure 6. We observed that rash was consistently associated with slower progression (HR: 0.78, CI [0.70, 0.87]), while hepatitis was associated with faster progression (HR: 1.43, CI [1.26, 1.62]). The results from the previous section indicated that several known prognostic markers were associated with the risk of rash, most notably the neutrophil-to-lymphocyte ratio (NLR). To disentangle the observed effect of irAEs on PFS from the general prognostic state at baseline, we conducted a bivariate two-stage meta-analysis by including irAE and the recently developed Real World Prognostic score (ROPRO) [25] as covariates in the Cox regression. ROPRO integrates clinical routine variables to develop a high-quality prognostic score for predicting overall survival in cancer patients. Interestingly, our analysis reveals that irAEs and ROPRO are independent predictors of PFS. While higher ROPRO values are associated with an increased hazard of progression, rash and hepatitis remain linked to slower and faster progression, respectively (see figure 6 and online supplemental figure 6).

**Figure 6:**
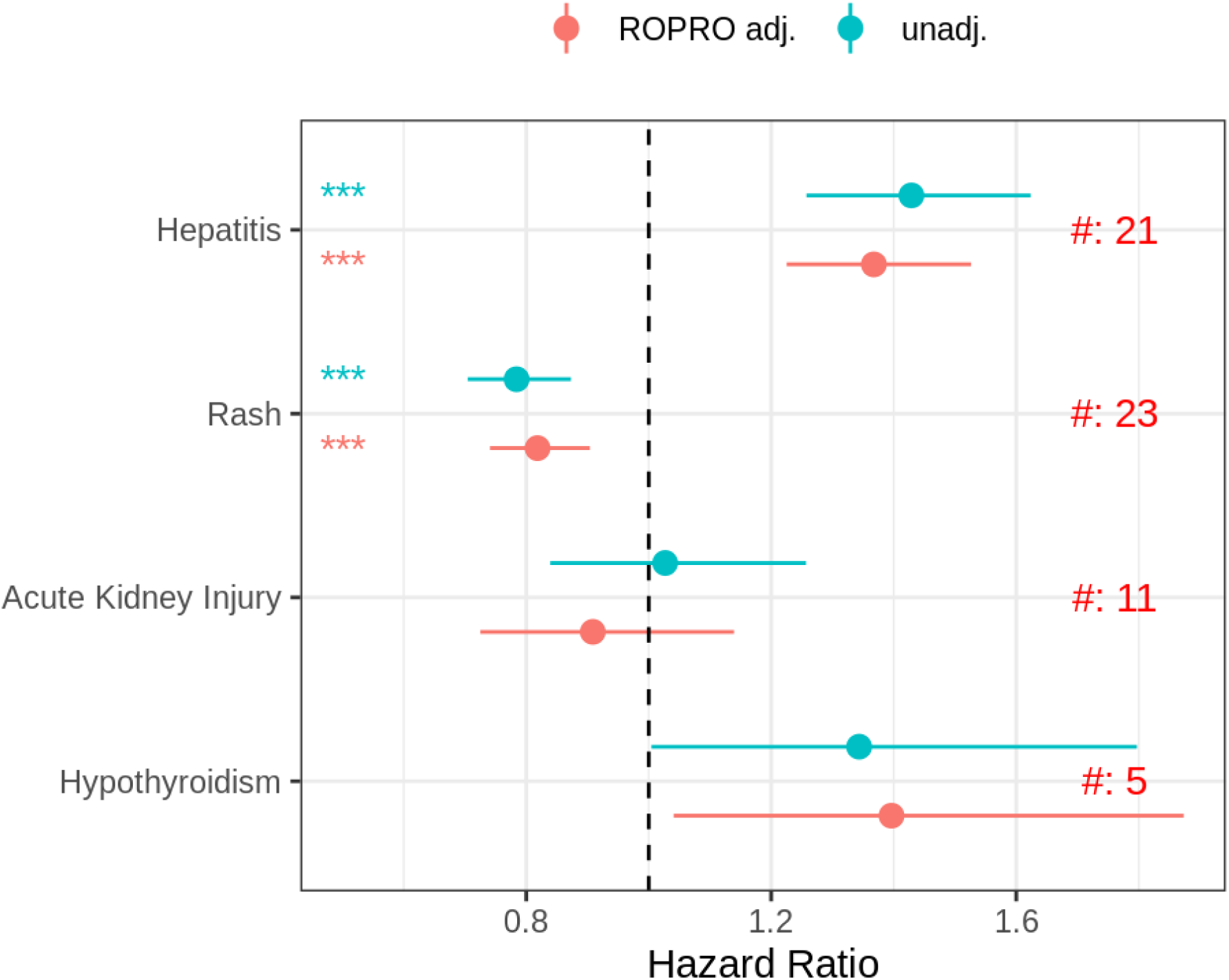
Association of irAEs with PFS, both with and without adjustment for ROPRO (Real World Prognostic score introduced in [25]). Hazard ratios (HR) and 95% confidence intervals (CI) were obtained through multivariate two-stage meta-analysis treating PFS as the time-to-event endpoint and irAE as covariate, with ROPRO as a potential control covariate. #[number] indicates the number of studies included in the meta-analysis. ** indicates results with a p-value <0.01, and *** indicates results with a p-value <0.001.

## Discussion

To the best of our knowledge, this analysis of 27 Roche-sponsored trials represents the first systematic investigation of irAEs associated with non-standard CITs in early-phase trials for advanced solid tumors. Understanding these adverse events is crucial for optimizing treatment strategies and improving patient outcomes. As expected, the incidence patterns of irAEs differ among CIT molecules and often align with anticipated trends based on their modes of action. Reassuringly, our incidence rates for treatments targeting the PD(L)1 axis are consistent with those reported in large anti-PD(L)1 trials. The findings on incidence proportions, severity, and onset can serve as a valuable resource for clinicians and drug developers to assess and compare the risk profiles of newly investigated immunotherapies, as well as to refine treatment regimens and the design of future trials.

Our analysis of risk factors is complicated by the significant heterogeneity among the included studies, particularly regarding molecule classes, study designs (e.g., varying dosing regimens or schedule of assessment), included indications, and concomitant treatments. This stands in stark contrast to similar investigations conducted in large phase 3 trials for standard checkpoint inhibitor treatments. To manage the differences across studies and avoid reporting associations that are only valid in one study, we used a robust statistical approach based on meta-analysis. This approach helped us find reliable associations that are valid pan-immunotherapy. Liver metastases correlate positively with hepatitis but appear protective against rash. Elevated liver enzyme levels are linked to hepatitis, higher ferritin levels are associated with an increased risk of acute kidney injury, and higher myeloid cell counts and elevated CRP levels seem to offer protection against rash. Notably, several of these markers were also identified as the strongest risk factors in a recent systematic review of atezolizumab trials [4]. Interestingly, we found moderate evidence suggesting that prior CPI exposure is associated with a reduced risk of hepatitis and hypothyroidism. Although the evidence is moderate, with upper confidence intervals nearing one, this unexpected finding lacks a complete explanation. One possible explanation could be selection bias; patients previously exposed to CPIs who were at risk for irAEs may have been excluded from further analysis. For instance, a patient with pre-existing hypothyroidism might be on long-term thyroxine therapy, reducing their likelihood of experiencing a subsequent hypothyroidism event.

Several recent studies involving large trials of standard CPIs have reported promising associations between genetic features and irAE risk. Consequently, we explored polygenic scores as potential risk factors. However, the observed effect sizes were small, and their predictive utility was limited in this early-phase trial setting, likely due to the heterogeneity of our study population, influenced by varying diseases and cancer types, as exemplified by our analysis of liver enzymes and corresponding PGSs.

It was hypothesized that variations in the TME might explain the risk of irAEs. However, our analysis of the TME using RNA sequencing data did not reveal any such association. This is in keeping with irAE development being more linked to features of systemic immunity relative to those of the TME.

We further evaluated irAEs as predictors of patient outcomes. Despite the heterogeneity of our cohort, we observed a consistent association between rash and improved PFS. Interestingly, our analysis suggests that this effect is independent of other baseline prognostic factors. These findings align with observations in the CPI setting [28,29], supporting the notion that skin toxicities may reflect a response at the tumor site and suggesting a broad immunotherapy benefit given the distinct mechanisms of action of CPIs and the drugs in this cohort. Conversely, other toxicities show no association with PFS or are prognostically unfavorable. Understanding the common features of skin toxicity and treatment response may offer insights for improved therapeutics.

Our study has several limitations that should be considered. Firstly, the safety follow-up duration across studies was both limited and variable, with a median follow-up time ranging from 2 to 15 months. This constraint restricted our investigation of the dynamics of irAEs to a 6-month period, thereby excluding the assessment of long-term and chronic events. Additionally, our analysis was confined to evaluating the time to the first occurrence of irAEs. Analyzing recurrent events, as well as the duration and resolution of these events, could have provided deeper insights into the overall safety profile of irAEs.

The studies included in our data mart exhibit significant heterogeneity across various aspects. For instance, exploring the impact of specific indications on the pattern of irAEs would have been insightful. However, the uneven distribution of indications across the studies could lead to misleading conclusions if irAE rates were compared naively by indication. This same issue limited our ability to identify context-specific risk factors. Nevertheless, in our risk factor analysis, we were able to investigate the impact of concomitant treatment with anti-PD-(L)1 therapies; however, the results indicate limited effect.

Our study included biomarkers of the main immune-cell populations, such as CD4 and CD8 T cells, B cells, and NK cells. Evaluating more fine-grained immune-cell subsets in the periphery would be of high interest; for example, CD4+ T effector memory cells have been reported to be predictive of irAEs. Similarly, a standardized cytokine panel would have been another interesting biomarker to investigate.

An interesting aspect of our evaluation of soluble biomarkers as predictors of irAE risk was the impact of including on-treatment samples. This is important because such data can guide patient monitoring. Our results indicate that on-treatment data significantly strengthens the association between liver enzymes and hepatitis. This finding is not surprising, as liver enzymes are used to diagnose hepatitis in the first place. However, we did not observe similar associations for other biomarkers. A notable limitation of our data mart was the substantial variation in on-treatment sampling and, consequently, data coverage across studies and biomarkers. Our analysis investigated the association of one risk factor at a time, with the aim of identifying a robust association. This can be seen as the first step in developing a prognostic or predictive score [30,31]. Development of such a score involves several steps of validation and might ultimately involve a combination of more than one factor. To our knowledge, no such validated score has been developed for irAEs, and admittedly, our work does not address this gap [7].

Finally, irAEs are not the only’unwanted’ immune responses associated with immunotherapies. Several of the molecules we investigated also encountered issues such as cytokine release syndrome and the development of anti-drug antibodies [32]. Our data mart provides an excellent starting point for exploring these topics further. Specifically, a systematic investigation of anti-drug antibodies in relation to non-standard CITs, with a particular focus on the role of the HLA region, offers an interesting avenue for future research [33,34].

Overall, our findings highlight the complexity of immune toxicities in the early-phase setting, emphasizing the importance of the CIT class, tumor burden, involved sites of metastases, and systemic immune state in the development of irAEs. These insights could guide future research and clinical strategies to mitigate risks and enhance therapeutic efficacy.

## Declarations

### Data availability statement

Due to Roche company policy and patient privacy reasons, access to individual patient-level data is restricted. For up-to-date details on Roche’s Global Policy on the Sharing of Clinical Information and how to request access to related clinical study documents, see here: https://go.roche.com/data_sharing.

### Author contribution

N.S. conceptualized and oversaw the study, with support from B.P.F., D.F.L., and V.C.S. Primary data analyses were conducted by N.S., D.F.L., and V.C.S.. The interpretation of results was carried out by N.S., B.P.F., D.F.L., V.C.S., R.M., V.K., D.H., and S.L.M. N.S. wrote the manuscript with input from B.P.F., D.F.L., and V.C.S. T.K.-T., P.S., and C.A. provided support with the analysis of genotyping data. The data mart effort was co-led by N.S., G.D.-N., and A.R. All authors reviewed and approved the final manuscript.

## Abbreviations

ALP: Alkaline phosphatase
ALT: Alanine aminotransferase
AST: Aspartate aminotransferase
BILI: Bilirubin
BsAbs: Bispecific antibodies
CI: 95% confidence interval
CIT: Cancer immunotherapy
CPI: Checkpoint inhibitor
CRP: C-reactive protein
CRS: Cytokine release syndrome
GGT: Gamma-glutamyl transpeptidase
HR: Hazard ratio
IL-2: Interleukin-2
irAE: Immune-related adverse event
LYMP: Lymphocytes
MONO: Monocytes
NEUT: Neutrophils
NLR: Neutrophil-to-lymphocyte ratio
PFS: Progression-free survival
PGS: Polygenic risk score
ROPRO: Real World Prognostic score
SLD: Sum of largest diameter of target lesions
TCB: T cell engaging bispecific antibody
TME: Tumor microenvironment
TSH: Thyroid-stimulating hormone
WBC: White blood cells

## Acknowledgements

The authors would like to thank the patients, their families, the participating study centers, and the members of the Enhanced Data Insights and Sharing network for their support in the curation and integration of the clinical data.

