## supplemental material for "Identification of Conserved Immune-Related Adverse Event Risk Factors and Clinical Outcomes in a Pan-Immunotherapy Data Mart"

#### Contents

**1 TME CD8 Inflammation Score and Gene Signatures** **1**

**2 Genotyping and Polygenic Risk Scores** **3**

**3 Additional Figures and Tables** **6**

**References** **27**

### 1 TME CD8 Inflammation Score and Gene Signatures

RNA-sequencing data were analyzed using *HTSeqGenie* in BioConductor [1] as follows: first, reads with low nucleotide qualities (70% of bases with quality <23) or matches to rRNA and adapter sequences were removed. The remaining reads were aligned to the human reference genome (human: GRCh38.p10, mouse: GRCm38.p5) using GSNAP (PMID:20147302, 27008021) version '2013-10-10-v2', allowing maximum of two mismatches per 75 base sequence (parameters: '-M 2 -n 10 -B 2 -i 1 -N 1 -w 200000 -E 1 –pairmax-rna=200000 –clip-overlap'). Transcript annotation was based on the Gencode genes data base (human: GENCODE 27, mouse: GENCODE M15). To quantify gene expression levels, the number of reads mapping unambiguously to the exons of each gene was calculated. In addition to nRPKM and raw counts, Counts Per Million (CPM) and Transcripts Per Million (TPM) expression values were calculated. Quality control was performed separately using the *helios* package.

The TME CD8 Inflammation Score was determined using an algorithm implemented in the package available at https://github.com/bedapub/cd8ippred, as described in [2]. The estimated probability of an 'inflamed phenotype' (`prob.inflamed`) was used as the TME CD8 Inflammation Score. Samples with `n_out_of_range` values greater than or equal to 3 were removed from the analysis. Gene signature scores, corresponding to the z-score of the Wilcoxon rank-sum test, were calculated using the *BioQC* package for the immune signatures provided in Table 1.

Table 1: Immune gene signatures

| Gene signature | Genes |
|---|---|
| Proliferating cell | MCM7, DTYMK, SKA2, SAE1, RFC2, GMPS, TMEM185A, MKI67, PCNA, STMN1, HMGB2 |
| IFNG | IDO1, CXCL10, CXCL9, HLA-DRA, STAT1, IFNG |
| Teffector | CD8A, GZMA, GZMB, IFNG, EOMES, PRF1, CXCL9, CXCL10, TBX21 |
| Cytotoxic T cell | TRGC2, GZMH, KLRD1, FGFBP2, GZMB, PRF1, SAMD3, MATK, GNLY, NKG7, GZMA |
| Exhausted CD8 T cell | PDCD1, HAVCR2, LAG3, ENTPD1, CD38, TOX |
| Regulatory T cell | FOXP3, CCR8, PMCH, CCR4, RTKN2, CTLA4 |
| B cell | CD19, MS4A1, TNFRSF13C, VPREB3, PAX5, CR2 |
| Myeloid cell | CSF3R, MS4A6A, MS4A7, MNDA, C5AR1, FCGR2A, C3AR1, FPR1, LILRB2, HDC, FCGR3B, CCL22 |
| MHC pathway | CALR, PSME2, TAP1, TAPBP, WSB1, PSMB9, PDIA3, PSMB8, IRF1, PSME1, CD74, PSME3 |

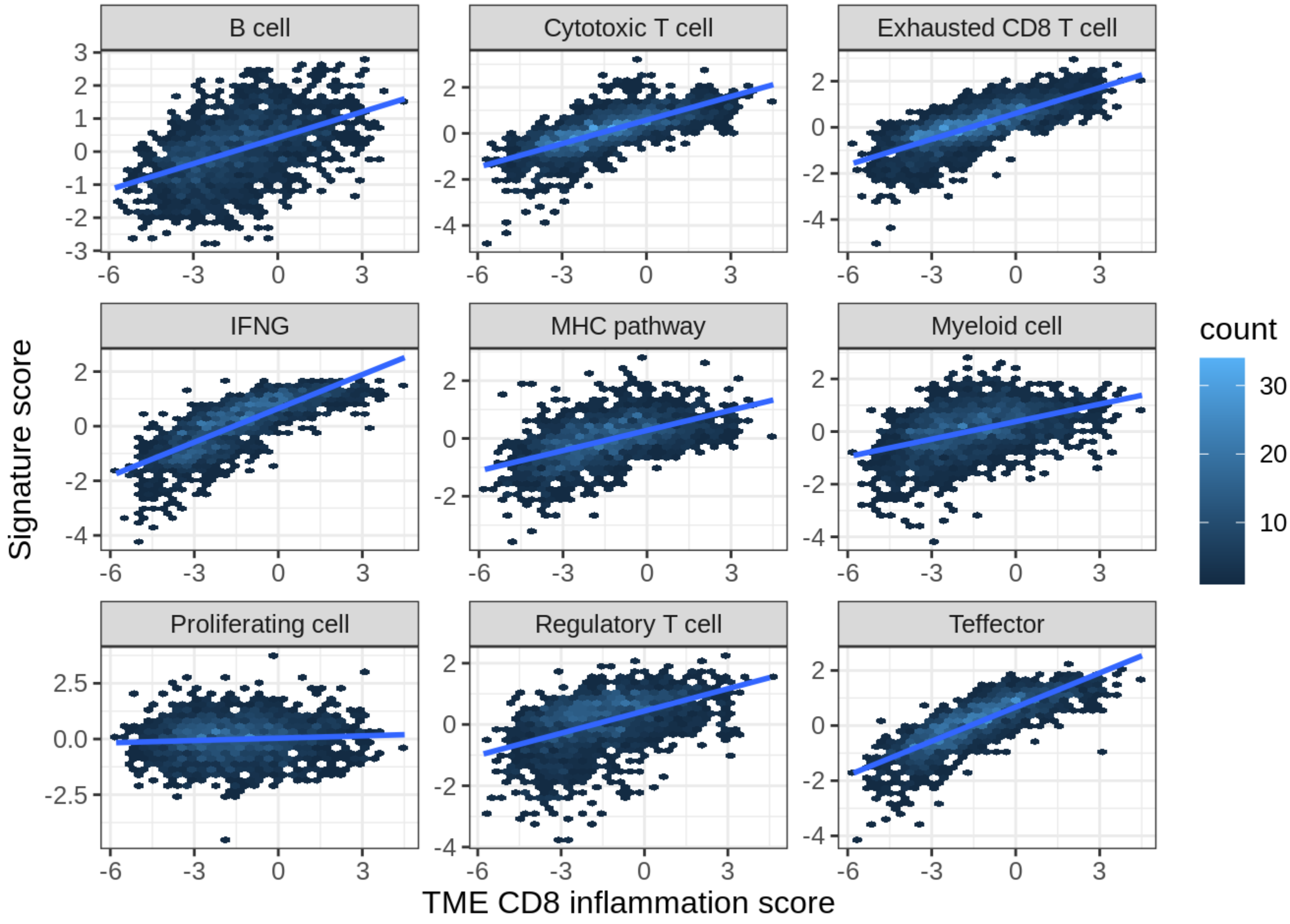


Figure 1: Scatter plot depicting gene signatures in relation to the predicted level of CD8+ T cell infiltration, represented by the TME CD8 inflammation score.

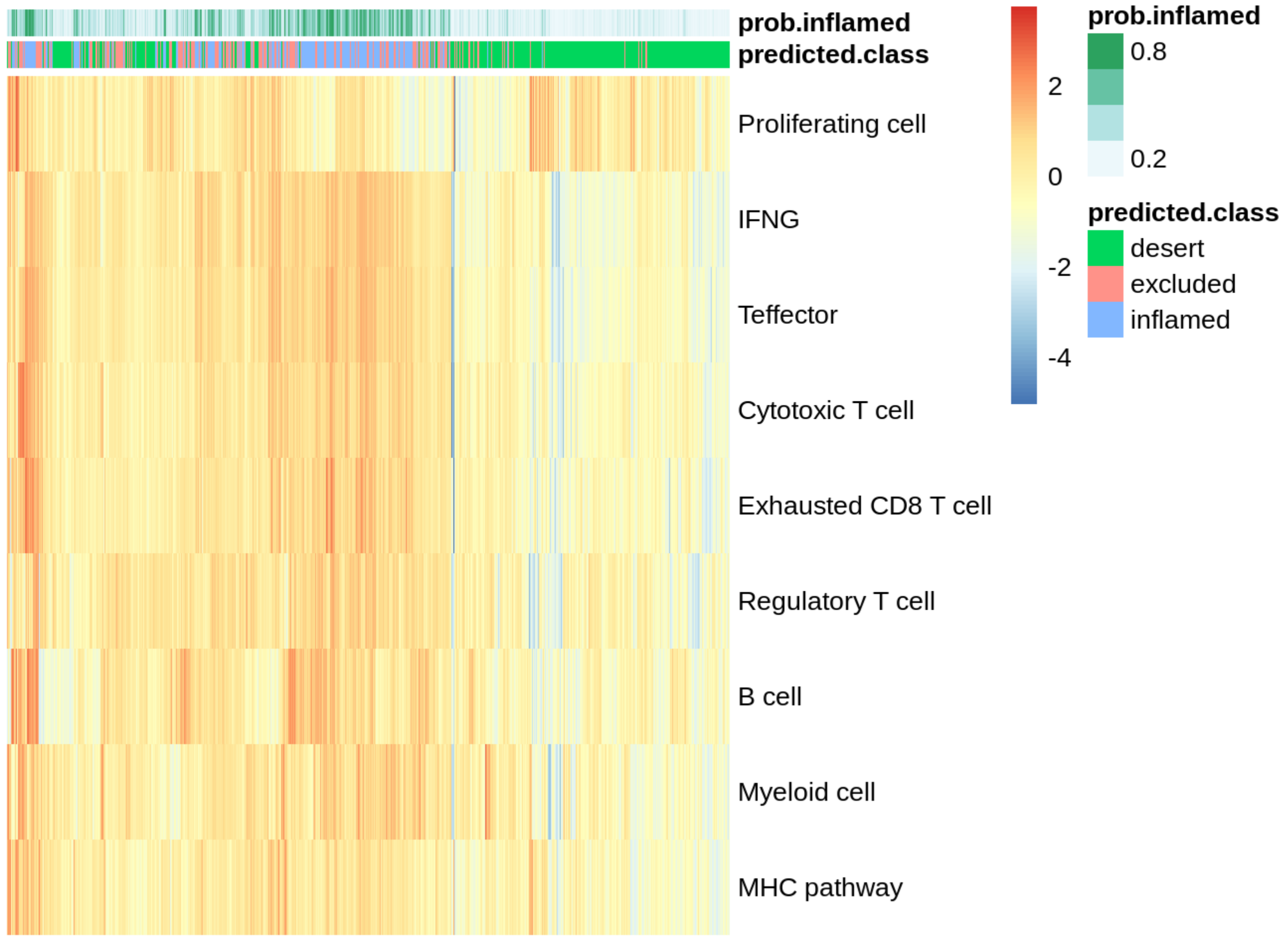


Figure 2: Summary of tumor microenvironment (TME) characteristics: Heatmap illustrating gene signatures, annotated by the predicted level of CD8+ T cell infiltration.

#### 2 Genotyping and Polygenic Risk Scores

A subset of 1929 patients was genotyped using the Illumina Global Screening Array, and imputation was performed as outlined in [3]. Principal component analysis was employed to infer ethnicity and X chromosome heterozygosity analysis was used to determine sex. Samples of non-European origin (221 out of 1929), and samples with discordance between reported and estimated gender (1 out of 1929) were excluded from downstream analysis.

Polygenic risk scores (PGS) were selected by linking irAE categories to disease-related terms and identifying all related subterms using the Experimental Factor Ontology (EFO) database (https://www.ebi.ac.uk/efo/). Subsequently, associated EFO subterms were extracted, and PGS were selected using references provided by the EBI Polygenic Risk Score Catalogue [4] based on the following criteria: 1) the PGS matches an EFO subterm, 2) the PGS is derived from data of European ancestry, and 3) if more than five PGS per reported trait were available, further selection was performed by choosing a maximum of five PGSs with the largest cohort sizes (see Table 2).

PGSs were then computed based on references provided by the EBI Polygenic Risk Score Catalogue (www.pgscatalog.org), excluding variants located in the major histocompatibility complex region. PGSs that had at least 75% of the variants captured and covered 75% of the sum of absolute weights were further considered for analysis.

Table 2: This table summarizes the polygenic risk scores selected for association analysis, including the following details: the irAE category, EFO search terms associated with the linked trait, EFO ID numbers used to identify all sub-traits according to the EFO database, the number of EFO subterms identified, the total number of PGS available in the PGS catalogue linked with the terms, the number of PGS derived from European ancestry (# of EUR PGS), the number of selected PGSs, and the selected PGS IDs.

| irAE category | EFO search terms | EFO ID Numbers | # of EFO subterms | Total PGS available | # of EUR PGS | # of selected PGSs | Selected PGS IDs |
|---|---|---|---|---|---|---|---|
| Hepatitis | liver disease, liver enzyme levels | EFO_0001421, EFO_0802260, EFO_0004533, EFO_0004736, EFO_0004532 | 61 | 27 | 24 | 24 | PGS004913, PGS000704, PGS004330, PGS000673, PGS001941, PGS002159, PGS003523, PGS004719, PGS004720, PGS004721, PGS004722, PGS000726, PGS000776, PGS004621, PGS001777, PGS004336, PGS000817, PGS001964, PGS002182, PGS003543, PGS000683, PGS001860, PGS002071, PGS001293 |
| Rash | atopic eczema, psoriasis, eczematoid dermatitis | EFO_0000274, EFO_0000676, HP_0000964 | 15 | 27 | 27 | 17 | PGS004587, PGS002755, PGS004903, PGS003486, PGS003459, PGS001773, PGS001871, PGS002083, PGS002344, PGS002416, PGS002465, PGS002514, PGS002563, PGS002612, PGS002661, PGS002710, PGS001313 |
| Acute kidney injury | chronic kidney disease | EFO_0003884 | 9 | 21 | 21 | 20 | PGS000728, PGS000859, PGS004224, PGS002757, PGS003988, PGS004004, PGS004016, PGS004030, PGS004045, PGS004058, PGS004074, PGS004088, PGS004101, PGS004112, PGS004128, PGS004142, PGS004158, PGS002237, PGS005090, PGS001272 |
| Hypothyroidism/ Hyperthyroidism | hypo-/hyperthyroidism, thyroid stimulating hormone | EFO_0009189, EFO_0004705, EFO_0004748 | 14 | 22 | 22 | 11 | PGS004446, PGS004516, PGS001043, PGS000759, PGS000761, PGS002766, PGS001816, PGS002024, PGS000820, PGS000965, PGS001181 |

Table 3: Overview of Included Studies: This table summarizes the analyzed studies, including details such as Molecule, Clinical Trial Number, STUDYID, AntiPDL1 Combination Status, Number of Subjects, Mean Age (with Range), and Percentage of Female Participants. Note: The BP40087 study comprises both single-agent and anti-PD-L1 combination therapy cohorts, which were treated as separate studies in our analysis.

| MOLECULE | Clinical Trial Number | STUDYID | AntiPDL1 Combination Status | Number of Subjects | Age (Mean, Range) | Percentage Female |
|---|---|---|---|---|---|---|
| TYRP1-TCB | NCT04551352 | BP42169 | | 20 | 59 (38,76) | 45 % |
| CEA-TCB | NCT02650713 | WP29945 | Yes | 228 | 56 (24,81) | 42 % |
| | NCT02324257 | BP29541 | | 149 | 59 (22,80) | 41 % |
| CEACAM5-TCB | NCT03539484 | BP40092 | | 26 | 60 (39,78) | 54 % |
| CEA-IL2v | NCT02004106 | BP28920 | | 113 | 61 (36,79) | 35 % |
| | NCT02350673 | BP29435 | Yes | 70 | 58 (28,79) | 51 % |
| FAP-IL2v | NCT03875079 | BP41054 | | 84 | 58 (21,86) | 45 % |
| | NCT02627274 | BP29842 | | 134 | 59 (35,79) | 40 % |
| | NCT03063762 | BP39365 | Yes | 69 | 58 (35,78) | 23 % |
| | NCT03386721 | BP40234 | Yes | 256 | 58 (19,80) | 43 % |
| FAP-41BBL | EudraCT: 2017-003961-83 | BP40087 | | 55 | 62 (31,84) | 47 % |
| | EudraCT: 2017-003961-83 | BP40087 | Yes | 94 | 58 (26,84) | 49 % |
| TLR7 | NCT04338685 | WP41377 | | 55 | 58 (37,77) | 24 % |
| BETi | NCT03068351 | NP39403 | | 24 | 65 (46,82) | 46 % |
| | NCT03292172 | NP39487 | Yes | 36 | 53 (34,72) | 100 % |
| | NCT03255096 | NP39461 | | 39 | 65 (28,81) | 51 % |
| CD40 | NCT02665416 | BP29889 | | 97 | 58 (23,80) | 56 % |
| | NCT02304393 | BP29392 | Yes | 140 | 57 (23,81) | 50 % |
| CSF1R | NCT02760797 | BP29427 | | 38 | 58 (35,78) | 55 % |
| | NCT01494688 | BP27772 | | 216 | 52 (18,82) | 65 % |
| | NCT02323191 | BP29428 | Yes | 221 | 61 (18,86) | 35 % |
| CD25 | NCT04642365 | BP42595 | Yes | 59 | 60 (25,80) | 44 % |
| | NCT04158583 | WP41188 | | 76 | 58 (34,80) | 57 % |
| OX40 | NCT02410512 | GO29674 | Yes | 298 | 59 (21,85) | 44 % |
| | NCT02219724 | GO29313 | | 172 | 58 (22,88) | 41 % |
| PD1/TIM3 | NCT03708328 | NP40435 | | 134 | 61 (31,81) | 37 % |
| PDL1 | NCT02814669 | BO30013 | Yes | 45 | 69 (41,85) | 0 % |
| | NCT01375842 | PCD4989G | Yes | 660 | 60 (20,89) | 47 % |

Table 4: Median follow-up time (in months) for Hepatitis across different studies, with 95% confidence intervals, calculated using the reverse Kaplan-Meier estimator.

| MOLECULE | STUDYID | Median | Lower 95% CI | Upper 95% CI |
|---|---|---|---|---|
| TYRP1-TCB | BP42169 | 4.57 | 2.40 | 10.68 |
| CEA-TCB | BP29541 | 2.99 | 2.37 | 3.91 |
| | WP29945 | 3.55 | 3.06 | 4.14 |
| CEACAM5-TCB | BP40092 | 2.89 | 1.71 | 3.15 |
| CEA-IL2v | BP28920 | 2.56 | 2.27 | 2.79 |
| | BP29435 | 2.37 | 1.94 | 3.71 |
| FAP-IL2v | BP29842 | 3.22 | 2.63 | 4.24 |
| | BP39365 | 14.92 | 2.46 | 24.90 |
| | BP40234 | 3.78 | 3.06 | 4.47 |
| | BP41054 | 3.71 | 2.40 | 6.54 |
| FAP-41BBL | BP40087_pdl1 | 4.47 | 4.01 | 5.65 |
| | BP40087_no_pdl1 | 2.63 | 2.40 | 4.07 |
| TLR7 | WP41377 | 2.60 | 2.17 | 3.52 |
| BETi | NP39403 | 2.07 | 1.48 | 3.06 |
| | NP39461 | 1.84 | 1.68 | 3.06 |
| | NP39487 | 2.17 | 2.10 | 3.06 |
| CD40 | BP29392 | 2.79 | 2.43 | 3.61 |
| | BP29889 | 4.27 | 3.32 | 4.70 |
| CSF1R | BP29428 | 2.83 | 2.40 | 3.09 |
| | BP27772 | 2.86 | 2.40 | 2.86 |
| | BP29427 | 2.27 | 1.74 | 3.12 |
| CD25 | BP42595 | 3.78 | 2.40 | 5.13 |
| | WP41188 | 2.40 | 2.37 | 2.43 |
| OX40 | GO29313 | 3.09 | 3.09 | 4.21 |
| | GO29674 | 3.09 | 3.09 | 3.32 |
| PD1/TIM3 | NP40435 | 3.32 | 2.40 | 3.78 |
| PDL1 | BO30013 | 5.09 | 3.35 | 6.54 |
| | PCD4989G | 4.37 | 3.75 | 4.47 |

### 3 Additional Figures and Tables

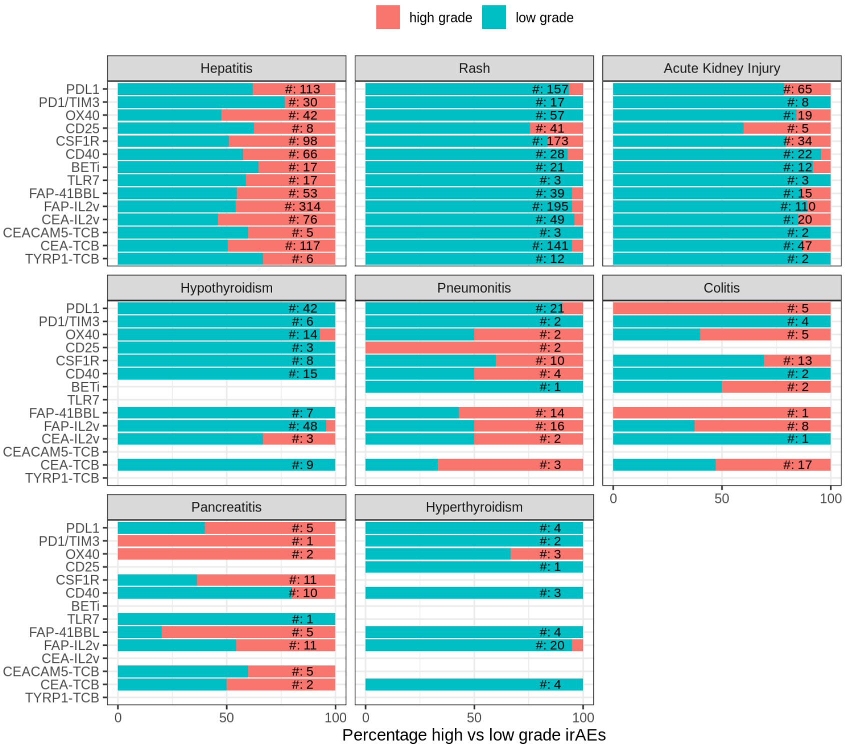


Figure 3: Percentage of high- vs. low-grade irAEs by CIT molecule. For each irAE category and CIT molecule, we compared the number of patients experiencing high-grade events (grades 3-5) to those experiencing low-grade events (grades 1-2). The NCI CTCAE criteria were utilized for grading the severity of irAEs.

Table 5: Association of tumor charactersitics with irAEs. Results of a two-stage meta-analysis: Hazard Ratios (HR), 95% Confidence Intervals (CI), p-values, heterogeneity statistic ($I^2$), and number of included studies (N.studies).

| irAE category | Tumor burden metric | HR | Lower 95% CI | Upper 95% CI | P.value | I2 [%] | N.studies |
|---|---|---|---|---|---|---|---|
| Hepatitis | with Liver Mets. | 1.99 | 1.45 | 2.74 | 0.00 | 69.57 | 18 |
| | with Lung Mets. | 0.93 | 0.83 | 1.04 | 0.17 | 0.00 | 21 |
| | # Metastases (log2) | 1.06 | 0.94 | 1.19 | 0.32 | 38.98 | 26 |
| | SLD (log2) | 1.09 | 0.98 | 1.21 | 0.12 | 38.00 | 22 |
| | TME CD8 inflam. score | 0.96 | 0.90 | 1.04 | 0.29 | 13.45 | 17 |
| Rash | with Liver Mets. | 0.75 | 0.62 | 0.91 | 0.01 | 14.92 | 18 |
| | with Lung Mets. | 0.88 | 0.72 | 1.08 | 0.20 | 27.83 | 22 |
| | # Metastases (log2) | 0.95 | 0.86 | 1.05 | 0.29 | 26.04 | 25 |
| | SLD (log2) | 0.80 | 0.74 | 0.87 | 0.00 | 0.00 | 22 |
| | TME CD8 inflam. score | 1.06 | 0.97 | 1.15 | 0.19 | 19.61 | 17 |
| Acute Kidney Injury | with Liver Mets. | 1.09 | 0.85 | 1.38 | 0.48 | 0.00 | 16 |
| | with Lung Mets. | 0.85 | 0.63 | 1.16 | 0.29 | 26.50 | 18 |
| | # Metastases (log2) | 1.09 | 0.97 | 1.24 | 0.14 | 0.00 | 19 |
| | SLD (log2) | 1.08 | 0.97 | 1.20 | 0.16 | 0.00 | 18 |
| | TME CD8 inflam. score | 1.02 | 0.92 | 1.14 | 0.64 | 0.00 | 10 |
| Hypothyroidism | with Liver Mets. | 1.05 | 0.88 | 1.25 | 0.53 | 0.00 | 8 |
| | with Lung Mets. | 1.10 | 0.82 | 1.48 | 0.47 | 0.00 | 9 |
| | # Metastases (log2) | 1.26 | 1.06 | 1.50 | 0.01 | 0.00 | 10 |
| | SLD (log2) | 0.94 | 0.78 | 1.12 | 0.44 | 0.00 | 9 |
| | TME CD8 inflam. score | 0.90 | 0.73 | 1.10 | 0.23 | 3.46 | 6 |

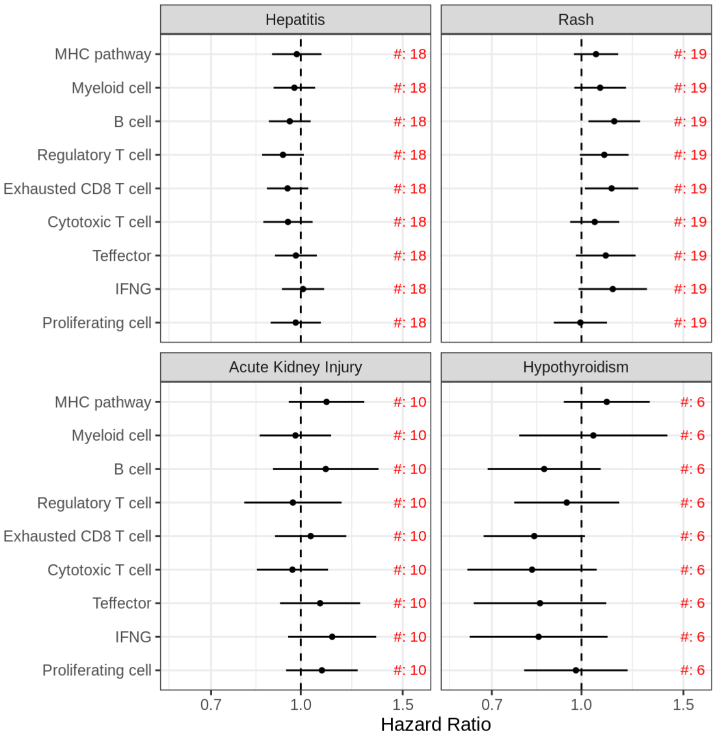


Figure 4: Association of tumor microenvironment (TME) gene signatures with irAEs. Results of a two-stage meta-analysis. #[number] indicates the number of studies included in the meta-analysis. ** indicates results with a p-value $< 0.01$, and *** indicates results with a p-value $< 0.001$.

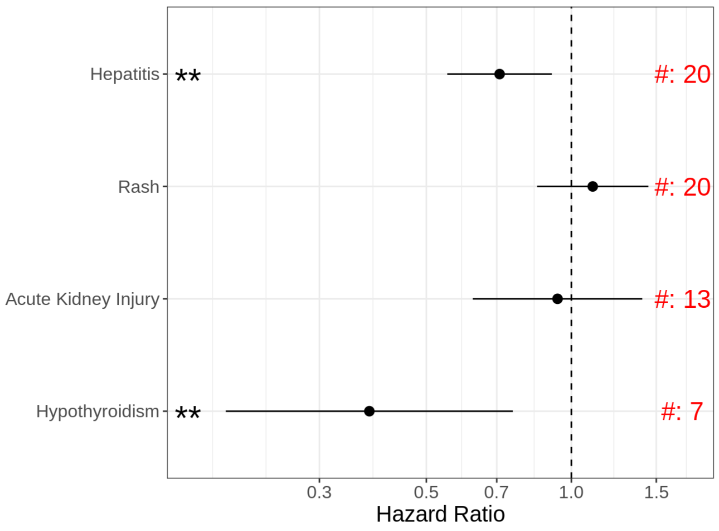


Figure 5: Association of previous CPI treatment with irAEs. Results of a two-stage meta-analysis. #[number] indicates the number of studies included in the meta-analysis. ** indicates results with a p-value $< 0.01$, and *** indicates results with a p-value $< 0.001$.

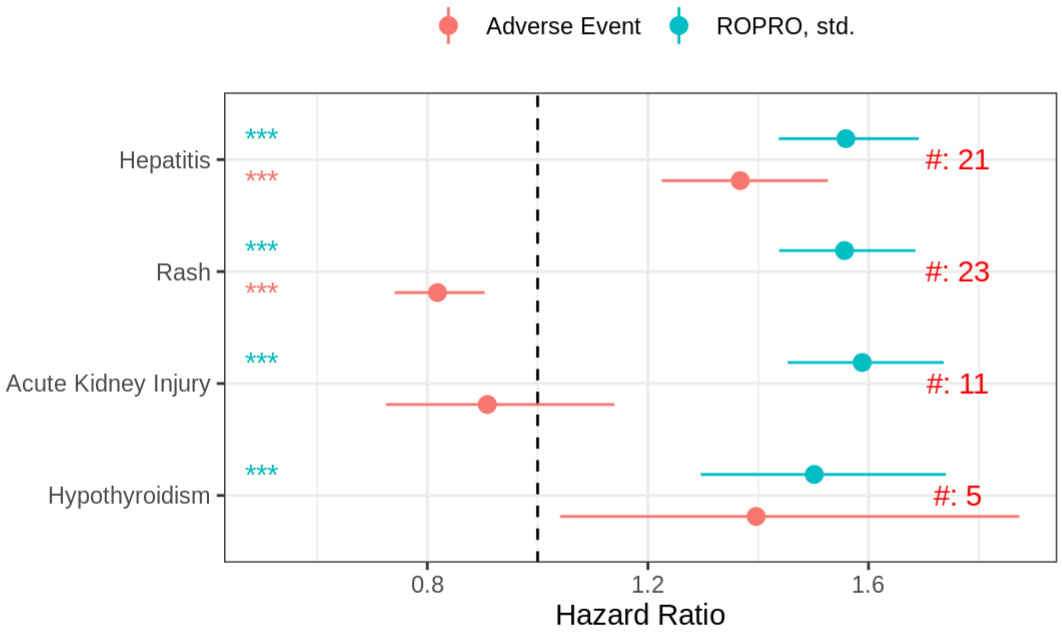


Figure 6: Association of irAEs and Real World Prognostic score (ROPRO) with PFS. Hazard ratios (HR) and 95% confidence intervals (CI) were obtained through bivariate two-stage meta-analysis treating PFS as the time-to-event endpoint and irAE and ROPRO as covariates. #[number] indicates the number of studies included in the meta-analysis. ** indicates results with a p-value $<0.01$, and *** indicates results with a p-value $<0.001$.

Table 6: Association of TME gene signatures with irAEs. Results of a two-stage meta-analysis. Hazard Ratios (HR), 95% Confidence Intervals (CI), p-values, heterogeneity statistic ($I^2$), and number of included studies (N.studies).

| irAE category | Gene signature | HR | Lower 95% CI | Upper 95% CI | P.value | I2 [%] | N.studies |
|---|---|---|---|---|---|---|---|
| Hepatitis | Proliferating cell | 0.98 | 0.89 | 1.08 | 0.69 | 25.19 | 18 |
| | IFNG | 1.01 | 0.93 | 1.10 | 0.84 | 0.00 | 18 |
| | Teffector | 0.98 | 0.90 | 1.07 | 0.65 | 0.00 | 18 |
| | Cytotoxic T cell | 0.95 | 0.86 | 1.05 | 0.31 | 20.43 | 18 |
| | Exhausted CD8 T cell | 0.95 | 0.87 | 1.03 | 0.21 | 0.00 | 18 |
| | Regulatory T cell | 0.93 | 0.86 | 1.01 | 0.09 | 0.01 | 18 |
| | B cell | 0.96 | 0.88 | 1.04 | 0.30 | 0.00 | 18 |
| | Myeloid cell | 0.97 | 0.90 | 1.06 | 0.54 | 0.00 | 18 |
| | MHC pathway | 0.98 | 0.89 | 1.09 | 0.75 | 14.09 | 18 |
| Rash | Proliferating cell | 1.00 | 0.90 | 1.11 | 0.93 | 20.12 | 19 |
| | IFNG | 1.13 | 0.99 | 1.30 | 0.07 | 36.64 | 19 |
| | Teffector | 1.10 | 0.98 | 1.24 | 0.11 | 25.01 | 19 |
| | Cytotoxic T cell | 1.05 | 0.96 | 1.16 | 0.29 | 2.80 | 19 |
| | Exhausted CD8 T cell | 1.13 | 1.01 | 1.25 | 0.03 | 15.00 | 19 |
| | Regulatory T cell | 1.09 | 0.99 | 1.21 | 0.07 | 0.00 | 19 |
| | B cell | 1.14 | 1.03 | 1.26 | 0.01 | 11.44 | 19 |
| | Myeloid cell | 1.08 | 0.97 | 1.19 | 0.16 | 8.84 | 19 |
| | MHC pathway | 1.06 | 0.97 | 1.16 | 0.20 | 0.01 | 19 |
| Acute Kidney Injury | Proliferating cell | 1.09 | 0.94 | 1.25 | 0.25 | 0.00 | 10 |
| | IFNG | 1.13 | 0.95 | 1.35 | 0.16 | 17.42 | 10 |
| | Teffector | 1.08 | 0.92 | 1.27 | 0.34 | 8.04 | 10 |
| | Cytotoxic T cell | 0.97 | 0.84 | 1.11 | 0.65 | 0.63 | 10 |
| | Exhausted CD8 T cell | 1.04 | 0.90 | 1.20 | 0.59 | 0.00 | 10 |
| | Regulatory T cell | 0.97 | 0.80 | 1.18 | 0.75 | 37.77 | 10 |
| | B cell | 1.10 | 0.90 | 1.36 | 0.35 | 46.61 | 10 |
| | Myeloid cell | 0.98 | 0.85 | 1.13 | 0.77 | 0.01 | 10 |
| | MHC pathway | 1.11 | 0.95 | 1.29 | 0.18 | 1.63 | 10 |
| Hypothyroidism | Proliferating cell | 0.98 | 0.80 | 1.20 | 0.83 | 0.01 | 6 |
| | IFNG | 0.84 | 0.64 | 1.11 | 0.22 | 26.70 | 6 |
| | Teffector | 0.85 | 0.65 | 1.10 | 0.22 | 23.81 | 6 |
| | Cytotoxic T cell | 0.82 | 0.64 | 1.06 | 0.13 | 32.02 | 6 |
| | Exhausted CD8 T cell | 0.83 | 0.68 | 1.01 | 0.07 | 0.00 | 6 |
| | Regulatory T cell | 0.94 | 0.77 | 1.16 | 0.58 | 0.00 | 6 |
| | B cell | 0.86 | 0.69 | 1.08 | 0.20 | 0.00 | 6 |
| | Myeloid cell | 1.05 | 0.78 | 1.41 | 0.75 | 37.32 | 6 |
| | MHC pathway | 1.11 | 0.93 | 1.31 | 0.25 | 0.00 | 6 |

Table 7: Association of previous CPI treatment with irAEs. Results of a two-stage meta-analysis. Hazard Ratios (HR), 95% Confidence Intervals (CI), p-values, heterogeneity statistic ($I^2$), and number of included studies (N.studies).

| irAE category | HR | Lower 95% CI | Upper 95% CI | P.value | I2 [%] | N.studies |
|---|---|---|---|---|---|---|
| Hepatitis | 0.71 | 0.55 | 0.91 | 0.01 | 26.73 | 20 |
| Rash | 1.11 | 0.85 | 1.44 | 0.45 | 36.53 | 20 |
| Acute Kidney Injury | 0.94 | 0.62 | 1.40 | 0.75 | 24.40 | 13 |
| Hypothyroidism | 0.38 | 0.19 | 0.76 | 0.01 | 0.00 | 7 |

Table 8: Association of soluble biomarkers at baseline with Hepatitis. Results of a two-stage meta-analysis: Hazard Ratios (HR), 95% Confidence Intervals (CI), p-values, heterogeneity statistic ($I^2$), and number of included studies (N.studies).

| | Hepatitis | | | | | |
|---|---|---|---|---|---|---|
| | HR | Lower 95% CI | Upper 95% CI | P.value | I2 [%] | N.studies |
| WBC | 0.95 | 0.86 | 1.06 | 0.38 | 44.91 | 22 |
| NEUT | 0.97 | 0.88 | 1.06 | 0.49 | 34.87 | 22 |
| MONO | 1.04 | 0.97 | 1.12 | 0.22 | 0.00 | 22 |
| LYM | 0.97 | 0.90 | 1.04 | 0.38 | 2.27 | 22 |
| NLR | 1.02 | 0.93 | 1.13 | 0.64 | 33.92 | 22 |
| B cells [%] | 1.03 | 0.95 | 1.11 | 0.50 | 0.00 | 16 |
| T cells [%] | 0.97 | 0.90 | 1.05 | 0.48 | 0.00 | 16 |
| CD4+ [% T cells] | 1.00 | 0.92 | 1.08 | 0.95 | 0.00 | 16 |
| CD8+ [% T cells] | 0.98 | 0.90 | 1.07 | 0.66 | 6.97 | 16 |
| NK cells [%] | 1.03 | 0.94 | 1.13 | 0.52 | 15.75 | 15 |
| AST | 1.56 | 1.38 | 1.76 | 0.00 | 64.90 | 21 |
| ALT | 1.36 | 1.23 | 1.50 | 0.00 | 40.57 | 21 |
| ALP | 1.40 | 1.23 | 1.59 | 0.00 | 69.08 | 20 |
| BILI | 1.18 | 1.09 | 1.28 | 0.00 | 10.83 | 21 |
| GGT | 1.34 | 1.15 | 1.56 | 0.00 | 68.50 | 17 |
| CRP | 1.06 | 0.94 | 1.19 | 0.32 | 41.59 | 19 |
| FERRITIN | 1.13 | 0.99 | 1.30 | 0.08 | 50.48 | 15 |
| TSH | 1.02 | 0.92 | 1.13 | 0.76 | 0.00 | 14 |

Table 9: Association of soluble biomarkers at baseline with Rash. Results of a two-stage meta-analysis.

| | Rash | | | | | |
|---|---|---|---|---|---|---|
| | HR | Lower 95% CI | Upper 95% CI | P.value | I2 [%] | N.studies |
| WBC | 0.90 | 0.84 | 0.96 | 0.00 | 1.83 | 22 |
| NEUT | 0.91 | 0.83 | 0.99 | 0.03 | 29.88 | 22 |
| MONO | 0.90 | 0.84 | 0.97 | 0.00 | 2.42 | 22 |
| LYM | 0.99 | 0.91 | 1.08 | 0.89 | 20.97 | 22 |
| NLR | 0.91 | 0.82 | 1.00 | 0.05 | 38.33 | 21 |
| B cells [%] | 1.09 | 1.00 | 1.19 | 0.05 | 13.84 | 16 |
| T cells [%] | 0.92 | 0.85 | 1.00 | 0.05 | 0.40 | 17 |
| CD4+ [% T cells] | 1.06 | 0.95 | 1.17 | 0.30 | 30.66 | 17 |
| CD8+ [% T cells] | 0.90 | 0.82 | 0.98 | 0.02 | 15.48 | 17 |
| NK cells [%] | 1.07 | 0.98 | 1.16 | 0.15 | 4.61 | 15 |
| AST | 0.92 | 0.85 | 1.00 | 0.05 | 13.71 | 21 |
| ALT | 1.01 | 0.93 | 1.11 | 0.76 | 29.99 | 22 |
| ALP | 0.85 | 0.77 | 0.94 | 0.00 | 28.71 | 21 |
| BILI | 1.05 | 0.98 | 1.12 | 0.19 | 0.00 | 22 |
| GGT | 0.87 | 0.80 | 0.94 | 0.00 | 0.00 | 17 |
| CRP | 0.82 | 0.73 | 0.91 | 0.00 | 44.24 | 18 |
| FERRITIN | 0.95 | 0.87 | 1.03 | 0.20 | 0.01 | 14 |
| TSH | 0.98 | 0.88 | 1.09 | 0.71 | 7.72 | 12 |

Table 10: Association of soluble biomarkers at baseline with Acute kidney injury. Results of a two-stage meta-analysis.

| | Acute kidney injury | | | | | |
|---|---|---|---|---|---|---|
| | HR | Lower 95% CI | Upper 95% CI | P.value | I2 [%] | N.studies |
| WBC | 1.21 | 1.07 | 1.36 | 0.00 | 0.00 | 16 |
| NEUT | 1.15 | 1.02 | 1.30 | 0.02 | 0.00 | 16 |
| MONO | 1.23 | 1.04 | 1.45 | 0.02 | 34.61 | 16 |
| LYM | 1.01 | 0.88 | 1.17 | 0.85 | 19.65 | 16 |
| NLR | 1.12 | 0.99 | 1.26 | 0.07 | 0.00 | 16 |
| B cells [%] | 0.89 | 0.76 | 1.05 | 0.17 | 33.98 | 11 |
| T cells [%] | 0.97 | 0.85 | 1.10 | 0.60 | 0.00 | 11 |
| CD4+ [% T cells] | 0.92 | 0.81 | 1.04 | 0.18 | 1.43 | 11 |
| CD8+ [% T cells] | 1.09 | 0.93 | 1.28 | 0.29 | 31.55 | 11 |
| NK cells [%] | 1.10 | 0.96 | 1.25 | 0.18 | 0.00 | 11 |
| AST | 1.00 | 0.88 | 1.13 | 0.96 | 0.00 | 16 |
| ALT | 0.93 | 0.82 | 1.05 | 0.22 | 0.00 | 16 |
| ALP | 1.09 | 0.97 | 1.23 | 0.15 | 0.00 | 15 |
| BILI | 0.95 | 0.85 | 1.07 | 0.43 | 3.10 | 16 |
| GGT | 1.06 | 0.93 | 1.22 | 0.37 | 0.00 | 13 |
| CRP | 1.21 | 0.99 | 1.49 | 0.07 | 49.56 | 13 |
| FERRITIN | 1.38 | 1.19 | 1.60 | 0.00 | 1.22 | 11 |
| TSH | 0.82 | 0.67 | 0.99 | 0.04 | 0.01 | 6 |

Table 11: Association of soluble biomarkers at baseline with Hypothyroidism. Results of a two-stage meta-analysis.

| | Hypothyroidism | | | | | |
|---|---|---|---|---|---|---|
| | HR | Lower 95% CI | Upper 95% CI | P.value | I2 [%] | N.studies |
| WBC | 0.88 | 0.72 | 1.07 | 0.20 | 0.00 | 8 |
| NEUT | 0.91 | 0.72 | 1.14 | 0.40 | 0.00 | 8 |
| MONO | 0.83 | 0.67 | 1.03 | 0.09 | 0.00 | 8 |
| LYM | 0.78 | 0.64 | 0.96 | 0.02 | 0.00 | 8 |
| NLR | 1.13 | 0.92 | 1.39 | 0.23 | 0.00 | 8 |
| B cells [%] | 1.10 | 0.90 | 1.36 | 0.35 | 0.01 | 6 |
| T cells [%] | 0.83 | 0.68 | 1.01 | 0.06 | 0.00 | 6 |
| CD4+ [% T cells] | 0.81 | 0.67 | 0.98 | 0.03 | 0.00 | 6 |
| CD8+ [% T cells] | 1.00 | 0.81 | 1.23 | 1.00 | 0.00 | 6 |
| NK cells [%] | 1.22 | 0.99 | 1.51 | 0.07 | 0.00 | 6 |
| AST | 1.00 | 0.82 | 1.23 | 0.97 | 0.00 | 8 |
| ALT | 0.94 | 0.77 | 1.14 | 0.51 | 0.00 | 8 |
| ALP | 0.97 | 0.79 | 1.18 | 0.75 | 1.34 | 8 |
| BILI | 0.91 | 0.74 | 1.11 | 0.34 | 19.88 | 8 |
| GGT | 0.94 | 0.71 | 1.23 | 0.63 | 0.00 | 6 |
| CRP | 0.93 | 0.75 | 1.15 | 0.51 | 0.01 | 6 |
| FERRITIN | 0.95 | 0.77 | 1.18 | 0.64 | 0.00 | 6 |
| TSH | 1.61 | 1.36 | 1.91 | 0.00 | 0.00 | 4 |

Table 12: Association of soluble biomarkers including on-treatment samples with Hepatitis. Results of a two-stage meta-analysis: Hazard Ratios (HR), 95% Confidence Intervals (CI), p-values, heterogeneity statistic ($I^2$), number of included studies (N.studies), and percentage coverage of on-treatment biomarker data.

| | Hepatitis | | | | | | |
|---|---|---|---|---|---|---|---|
| | HR | Lower 95% CI | Upper 95% CI | P.value | I2 [%] | N.studies | On-treatment coverage [%] |
| WBC | 0.95 | 0.86 | 1.05 | 0.29 | 50.83 | 22 | 48.77 |
| NEUT | 0.96 | 0.86 | 1.08 | 0.53 | 55.44 | 22 | 53.28 |
| MONO | 0.94 | 0.88 | 1.00 | 0.05 | 21.91 | 22 | 53.33 |
| LYM | 0.93 | 0.89 | 0.98 | 0.00 | 0.00 | 22 | 53.17 |
| NLR | 1.06 | 0.98 | 1.13 | 0.13 | 26.45 | 22 | 51.71 |
| B cells [%] | 1.01 | 0.95 | 1.08 | 0.67 | 0.01 | 16 | 36.78 |
| T cells [%] | 0.98 | 0.92 | 1.05 | 0.55 | 13.20 | 16 | 36.73 |
| CD4+ [% T cells] | 1.01 | 0.94 | 1.07 | 0.87 | 0.00 | 16 | 36.70 |
| CD8+ [% T cells] | 1.00 | 0.93 | 1.08 | 0.90 | 10.10 | 16 | 36.78 |
| NK cells [%] | 1.00 | 0.94 | 1.07 | 0.97 | 0.00 | 15 | 34.96 |
| AST | 1.73 | 1.59 | 1.88 | 0.00 | 54.56 | 21 | 48.62 |
| ALT | 1.67 | 1.54 | 1.81 | 0.00 | 34.24 | 21 | 48.83 |
| ALP | 1.52 | 1.36 | 1.70 | 0.00 | 64.30 | 20 | 49.74 |
| BILI | 1.40 | 1.29 | 1.52 | 0.00 | 33.05 | 21 | 48.73 |
| GGT | 1.59 | 1.43 | 1.77 | 0.00 | 30.87 | 17 | 74.29 |
| CRP | 1.23 | 1.11 | 1.36 | 0.00 | 0.00 | 19 | 59.76 |
| FERRITIN | 1.27 | 1.13 | 1.41 | 0.00 | 24.41 | 15 | 56.54 |
| TSH | 1.04 | 0.94 | 1.15 | 0.46 | 0.00 | 14 | 11.34 |

Table 13: Association of soluble biomarkers including on-treatment samples with Rash. Results of a two-stage meta-analysis.

| | Rash | | | | | | |
|---|---|---|---|---|---|---|---|
| | HR | Lower 95% CI | Upper 95% CI | P.value | I2 [%] | N.studies | On-treatment coverage [%] |
| WBC | 0.89 | 0.84 | 0.95 | 0.00 | 0.00 | 22 | 50.21 |
| NEUT | 0.87 | 0.80 | 0.94 | 0.00 | 17.96 | 22 | 54.90 |
| MONO | 0.92 | 0.87 | 0.98 | 0.01 | 4.69 | 22 | 55.03 |
| LYM | 1.02 | 0.95 | 1.09 | 0.56 | 23.22 | 22 | 55.07 |
| NLR | 0.91 | 0.84 | 0.98 | 0.02 | 32.56 | 21 | 55.44 |
| B cells [%] | 1.03 | 0.96 | 1.11 | 0.36 | 0.00 | 16 | 40.39 |
| T cells [%] | 1.00 | 0.94 | 1.06 | 0.94 | 0.00 | 17 | 40.22 |
| CD4+ [% T cells] | 1.06 | 0.97 | 1.15 | 0.19 | 23.45 | 17 | 40.19 |
| CD8+ [% T cells] | 0.92 | 0.85 | 0.99 | 0.03 | 13.04 | 17 | 40.26 |
| NK cells [%] | 0.99 | 0.92 | 1.07 | 0.89 | 11.05 | 15 | 38.72 |
| AST | 0.98 | 0.93 | 1.05 | 0.60 | 1.62 | 21 | 50.80 |
| ALT | 1.07 | 1.00 | 1.14 | 0.05 | 6.61 | 22 | 49.80 |
| ALP | 0.88 | 0.81 | 0.96 | 0.00 | 29.38 | 21 | 50.90 |
| BILI | 1.03 | 0.96 | 1.10 | 0.43 | 8.90 | 22 | 49.78 |
| GGT | 0.90 | 0.83 | 0.98 | 0.01 | 0.00 | 17 | 77.13 |
| CRP | 0.84 | 0.74 | 0.95 | 0.01 | 46.14 | 18 | 63.85 |
| FERRITIN | 0.96 | 0.88 | 1.05 | 0.34 | 0.01 | 14 | 60.39 |
| TSH | 0.97 | 0.88 | 1.06 | 0.47 | 0.00 | 12 | 11.59 |

Table 14: Association of soluble biomarkers including on-treatment samples with Acute kidney injury. Results of a two-stage meta-analysis.

| | Acute kidney injury | | | | | | |
|---|---|---|---|---|---|---|---|
| | HR | Lower 95% CI | Upper 95% CI | P.value | I2 [%] | N.studies | On-treatment coverage [%] |
| WBC | 1.24 | 1.13 | 1.38 | 0.00 | 0.01 | 16 | 49.62 |
| NEUT | 1.22 | 1.09 | 1.36 | 0.00 | 0.00 | 16 | 54.70 |
| MONO | 1.12 | 0.99 | 1.28 | 0.07 | 36.75 | 16 | 54.79 |
| LYM | 0.93 | 0.85 | 1.01 | 0.09 | 0.00 | 16 | 54.83 |
| NLR | 1.19 | 1.09 | 1.30 | 0.00 | 0.01 | 16 | 53.81 |
| B cells [%] | 0.94 | 0.83 | 1.08 | 0.40 | 21.84 | 11 | 36.66 |
| T cells [%] | 0.95 | 0.85 | 1.05 | 0.30 | 2.69 | 11 | 36.65 |
| CD4+ [% T cells] | 0.92 | 0.83 | 1.02 | 0.13 | 0.00 | 11 | 36.65 |
| CD8+ [% T cells] | 1.08 | 0.96 | 1.20 | 0.20 | 0.00 | 11 | 36.65 |
| NK cells [%] | 1.10 | 0.98 | 1.22 | 0.10 | 0.00 | 11 | 36.71 |
| AST | 1.01 | 0.92 | 1.11 | 0.88 | 0.00 | 16 | 49.12 |
| ALT | 0.95 | 0.86 | 1.05 | 0.33 | 0.01 | 16 | 49.28 |
| ALP | 1.13 | 1.02 | 1.25 | 0.02 | 0.00 | 15 | 50.41 |
| BILI | 1.09 | 0.97 | 1.23 | 0.14 | 21.22 | 16 | 49.33 |
| GGT | 1.09 | 0.95 | 1.27 | 0.22 | 0.00 | 13 | 76.50 |
| CRP | 1.40 | 1.10 | 1.77 | 0.01 | 38.75 | 13 | 63.87 |
| FERRITIN | 1.44 | 1.24 | 1.68 | 0.00 | 0.00 | 11 | 60.07 |
| TSH | 0.82 | 0.68 | 0.99 | 0.04 | 0.00 | 6 | 15.70 |

Table 15: Association of soluble biomarkers including on-treatment samples with Hypothyroidism. Results of a two-stage meta-analysis.

| | Hypothyroidism | | | | | | |
|---|---|---|---|---|---|---|---|
| | HR | Lower 95% CI | Upper 95% CI | P.value | I2 [%] | N.studies | On-treatment coverage [%] |
| WBC | 0.89 | 0.73 | 1.07 | 0.22 | 0.00 | 8 | 47.97 |
| NEUT | 0.92 | 0.75 | 1.14 | 0.47 | 0.00 | 8 | 55.19 |
| MONO | 0.95 | 0.79 | 1.15 | 0.63 | 0.00 | 8 | 55.19 |
| LYM | 0.76 | 0.63 | 0.92 | 0.00 | 0.00 | 8 | 55.22 |
| NLR | 1.14 | 0.94 | 1.37 | 0.18 | 0.00 | 8 | 54.83 |
| B cells [%] | 1.13 | 0.92 | 1.37 | 0.24 | 0.00 | 6 | 18.12 |
| T cells [%] | 0.84 | 0.70 | 1.00 | 0.05 | 0.00 | 6 | 18.20 |
| CD4+ [% T cells] | 0.84 | 0.70 | 1.00 | 0.05 | 0.00 | 6 | 18.20 |
| CD8+ [% T cells] | 0.96 | 0.79 | 1.18 | 0.71 | 0.00 | 6 | 18.20 |
| NK cells [%] | 1.15 | 0.93 | 1.42 | 0.19 | 4.04 | 6 | 18.20 |
| AST | 0.94 | 0.80 | 1.10 | 0.41 | 0.92 | 8 | 47.39 |
| ALT | 0.91 | 0.77 | 1.07 | 0.25 | 0.00 | 8 | 47.67 |
| ALP | 0.95 | 0.83 | 1.09 | 0.46 | 0.00 | 8 | 47.53 |
| BILI | 0.86 | 0.73 | 1.02 | 0.08 | 0.00 | 8 | 47.81 |
| GGT | 0.86 | 0.65 | 1.14 | 0.29 | 0.00 | 6 | 88.38 |
| CRP | 0.93 | 0.72 | 1.20 | 0.59 | 0.00 | 6 | 66.72 |
| FERRITIN | 0.95 | 0.75 | 1.19 | 0.65 | 0.00 | 6 | 62.24 |
| TSH | 1.58 | 1.33 | 1.87 | 0.00 | 0.00 | 4 | 16.83 |

Table 16: Association with PGS linked to irAEs. Results of a two-stage meta-analysis. Hazard Ratios (HR), 95% Confidence Intervals (CI), p-values, heterogeneity statistic ($I^2$), and number of included studies (N.studies).

| irAE category | PGS | HR | Lower 95% CI | Upper 95% CI | P.value | I2 [%] | N.studies |
|---|---|---|---|---|---|---|---|
| Hepatitis | Gamma-glutamyl transferase PGS000817 | 1.00 | 0.89 | 1.12 | 0.96 | 26.29 | 17 |
| | Alcoholic cirrhosis PGS000704 | 1.01 | 0.92 | 1.11 | 0.84 | 0.00 | 17 |
| | Aspartate aminotransferase [U/L] PGS000673 | 1.02 | 0.93 | 1.12 | 0.65 | 0.00 | 17 |
| | Gamma-glutamyl transferase [U/L] PGS000683 | 1.03 | 0.90 | 1.18 | 0.62 | 41.22 | 17 |
| | Gamma-glutamyl transferase PGS003543 | 1.04 | 0.91 | 1.18 | 0.57 | 38.99 | 17 |
| | Cirrhosis PGS000776 | 1.04 | 0.95 | 1.14 | 0.40 | 0.00 | 17 |
| | Gamma-glutamyl transferase PGS001964 | 1.06 | 0.92 | 1.21 | 0.42 | 43.45 | 17 |
| | Aspartate aminotransferase level PGS003523 | 1.06 | 0.97 | 1.16 | 0.23 | 0.00 | 17 |
| | Gamma-glutamyl transferase PGS002182 | 1.06 | 0.93 | 1.21 | 0.40 | 39.47 | 17 |
| | Cirrhosis PGS000726 | 1.06 | 0.97 | 1.16 | 0.22 | 0.00 | 17 |
| | Aspartate aminotransferase level PGS002159 | 1.06 | 0.97 | 1.17 | 0.18 | 0.00 | 17 |
| | Aspartate aminotransferase level PGS001941 | 1.07 | 0.98 | 1.17 | 0.15 | 0.00 | 17 |
| | Other chronic nonalcoholic liver disease PGS002071 | 1.10 | 1.00 | 1.21 | 0.06 | 0.00 | 17 |
| | Other chronic nonalcoholic liver disease PGS001860 | 1.11 | 1.01 | 1.22 | 0.03 | 0.00 | 17 |
| Rash | Psoriasis PGS002344 | 1.01 | 0.91 | 1.13 | 0.79 | 23.20 | 16 |
| | Psoriasis PGS002416 | 1.01 | 0.79 | 1.29 | 0.95 | 0.54 | 16 |
| | Psoriasis PGS002465 | 0.92 | 0.69 | 1.23 | 0.58 | 2.26 | 16 |
| | Psoriasis PGS002514 | 0.77 | 0.54 | 1.10 | 0.15 | 10.37 | 16 |
| | Psoriasis PGS002563 | 1.15 | 0.93 | 1.42 | 0.19 | 0.00 | 16 |
| | Psoriasis PGS002612 | 1.08 | 0.88 | 1.33 | 0.44 | 2.04 | 16 |
| | Psoriasis PGS002661 | 0.96 | 0.85 | 1.07 | 0.44 | 0.00 | 16 |
| | Psoriasis PGS002710 | 1.02 | 0.93 | 1.12 | 0.68 | 12.32 | 16 |
| | Atopic dermatitis PGS002755 | 0.89 | 0.66 | 1.20 | 0.44 | 41.15 | 16 |
| | Atopic eczema or atopic disease PGS003459 | 0.96 | 0.88 | 1.05 | 0.41 | 0.00 | 16 |
| | Atopic eczema PGS003486 | 0.96 | 0.87 | 1.05 | 0.33 | 0.00 | 16 |
| Acute Kidney Injury | Chronic kidney disease PGS002757 | 0.91 | 0.75 | 1.10 | 0.31 | 20.43 | 13 |
| | Chronic kidney disease (CKD) PGS004128 | 1.03 | 0.86 | 1.22 | 0.77 | 12.72 | 13 |
| | Chronic kidney disease (CKD) PGS004112 | 1.04 | 0.87 | 1.24 | 0.65 | 11.38 | 13 |
| | Chronic kidney disease (CKD) PGS004004 | 1.07 | 0.89 | 1.29 | 0.48 | 21.21 | 13 |
| | Chronic kidney disease PGS000728 | 1.08 | 0.83 | 1.41 | 0.55 | 0.00 | 13 |
| | Chronic kidney disease (CKD) PGS004088 | 1.09 | 0.90 | 1.31 | 0.40 | 16.31 | 13 |
| | Chronic kidney disease (stage 3 or greater) PGS002237 | 1.09 | 0.92 | 1.29 | 0.34 | 13.83 | 13 |
| | Chronic kidney disease (CKD) PGS004016 | 1.09 | 0.92 | 1.29 | 0.30 | 3.78 | 13 |
| | Chronic kidney disease (CKD) PGS004101 | 1.10 | 0.91 | 1.33 | 0.32 | 18.90 | 13 |
| | Chronic kidney disease (CKD) PGS003988 | 1.11 | 0.93 | 1.33 | 0.24 | 5.37 | 13 |
| | Chronic kidney disease PGS004224 | 1.11 | 0.91 | 1.36 | 0.29 | 26.79 | 13 |
| | Chronic kidney disease (CKD) PGS004058 | 1.12 | 0.95 | 1.32 | 0.19 | 0.00 | 13 |
| | Chronic kidney disease (CKD) PGS004074 | 1.14 | 0.95 | 1.37 | 0.16 | 12.61 | 13 |
| | Chronic kidney disease (CKD) PGS004045 | 1.15 | 0.95 | 1.38 | 0.14 | 15.28 | 13 |
| | Chronic kidney disease (CKD) PGS004158 | 1.16 | 0.97 | 1.38 | 0.10 | 5.64 | 13 |
| | Chronic kidney disease (CKD) PGS004030 | 1.16 | 0.96 | 1.42 | 0.13 | 24.40 | 13 |
| | Chronic kidney disease (CKD) PGS004142 | 1.18 | 0.98 | 1.42 | 0.08 | 18.88 | 13 |
| Hypothyroidism | Hypothyroidism (self-reported) PGS000820 | 1.06 | 0.81 | 1.38 | 0.68 | 0.00 | 7 |
| | Hypothyroidism PGS000761 | 1.09 | 0.81 | 1.45 | 0.58 | 13.44 | 7 |
| | Hypothyroidism PGS002024 | 1.09 | 0.73 | 1.64 | 0.68 | 48.58 | 7 |
| | Hypothyroidism PGS000759 | 1.10 | 0.80 | 1.53 | 0.56 | 33.46 | 7 |
| | Hypothyroidism PGS002766 | 1.11 | 0.74 | 1.65 | 0.61 | 48.56 | 7 |
| | Other hypothyroidism (time-to-event) PGS001181 | 1.13 | 0.86 | 1.49 | 0.38 | 0.00 | 7 |
| | Hypothyroidism/myxoedema PGS000965 | 1.13 | 0.85 | 1.52 | 0.40 | 8.03 | 7 |
| | Hypothyroidism PGS001816 | 1.20 | 0.74 | 1.95 | 0.46 | 63.03 | 7 |

Table 17: Variance explained by PGS developed for liver enzymes with corresponding protein measurements, stratified by liver metastasis status. Variance explained and 95% CI were obtained through univariate two-stage meta-analysis.

| PGS - Liver Enzyme | Liver Mets. Status | Variance Explained [%] | Lower 95% CI | Upper 95% CI | I2 [%] |
|---|---|---|---|---|---|
| ALP | with Liver mets. | 0.47 | 0.00 | 2.56 | 0.00 |
| | without Liver mets. | 2.48 | 0.69 | 5.36 | 29.32 |
| ALT | with Liver mets. | 0.41 | 0.00 | 1.85 | 0.00 |
| | without Liver mets. | 1.56 | 0.27 | 3.91 | 4.30 |
| AST | with Liver mets. | 1.10 | 0.00 | 4.43 | 0.34 |
| | without Liver mets. | 1.06 | 0.39 | 2.07 | 0.00 |
| GGT | with Liver mets. | 2.73 | 0.62 | 6.32 | 0.00 |
| | without Liver mets. | 3.13 | 1.09 | 6.23 | 0.00 |

Table 18: Association of irAEs with PFS, excluding patients with reported liver metastasis. Results of a two-stage meta-analysis. Hazard Ratios (HR), 95% Confidence Intervals (CI), and p-values.

| irAE category | Strata | HR | Lower 95% CI | Upper 95% CI | P.value |
|---|---|---|---|---|---|
| Hepatitis | all subjects | 1.43 | 1.26 | 1.62 | 0.00 |
| | no liver mets | 1.28 | 1.11 | 1.47 | 0.00 |
| Rash | all subjects | 0.78 | 0.70 | 0.87 | 0.00 |
| | no liver mets | 0.80 | 0.70 | 0.92 | 0.00 |
| Acute Kidney Injury | all subjects | 1.03 | 0.84 | 1.26 | 0.80 |
| | no liver mets | 0.99 | 0.75 | 1.30 | 0.94 |
| Hypothyroidism | all subjects | 1.34 | 1.00 | 1.80 | 0.05 |
| | no liver mets | 1.54 | 1.12 | 2.11 | 0.01 |

Table 19: Association of irAEs with PFS either with or without adjustment for ROPRO (Real World Prognostic score). Hazard ratios (HR) and 95% confidence intervals (CI) were obtained through multivariate two-stage meta-analysis treating PFS as the time-to-event endpoint and irAE as covariate, with ROPRO as a potential control covariate.

| irAE category | Correction | HR | Lower 95% CI | Upper 95% CI | P.value | I2 [%] | N.studies |
|---|---|---|---|---|---|---|---|
| Hepatitis | unadj. | 1.43 | 1.26 | 1.62 | 0.00 | 35.89 | 21 |
| | ROPRO adj. | 1.37 | 1.23 | 1.53 | 0.00 | 16.06 | 21 |
| Rash | unadj. | 0.78 | 0.70 | 0.87 | 0.00 | 10.79 | 23 |
| | ROPRO adj. | 0.82 | 0.74 | 0.90 | 0.00 | 0.00 | 23 |
| Acute Kidney Injury | unadj. | 1.03 | 0.84 | 1.26 | 0.80 | 34.71 | 11 |
| | ROPRO adj. | 0.91 | 0.72 | 1.14 | 0.41 | 46.88 | 11 |
| Hypothyroidism | unadj. | 1.34 | 1.00 | 1.80 | 0.05 | 0.00 | 5 |
| | ROPRO adj. | 1.40 | 1.04 | 1.87 | 0.03 | 0.00 | 5 |

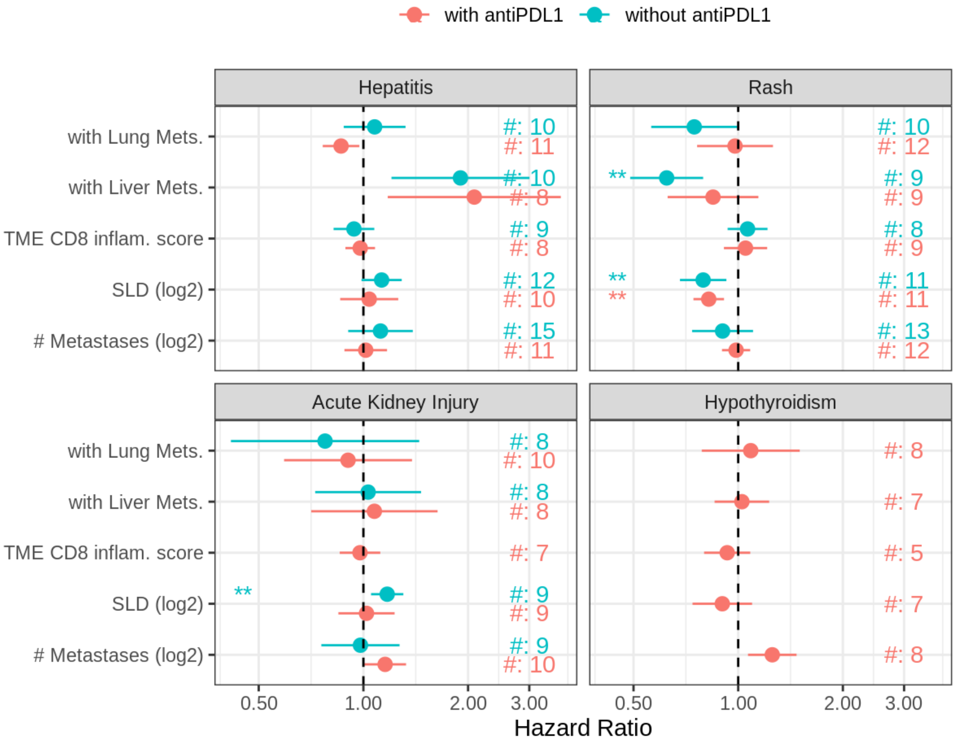


Figure 7: Association of tumor burden metrics with irAEs stratified by Anti-PD-L1 combination status. Hazard ratios (HR) and 95% confidence intervals (CI) were obtained through a confounder-adjusted two-stage meta-analysis. #[number] indicates the number of studies included in the meta-analysis. ** indicates results with a p-value <0.01, and *** indicates results with a p-value <0.001.

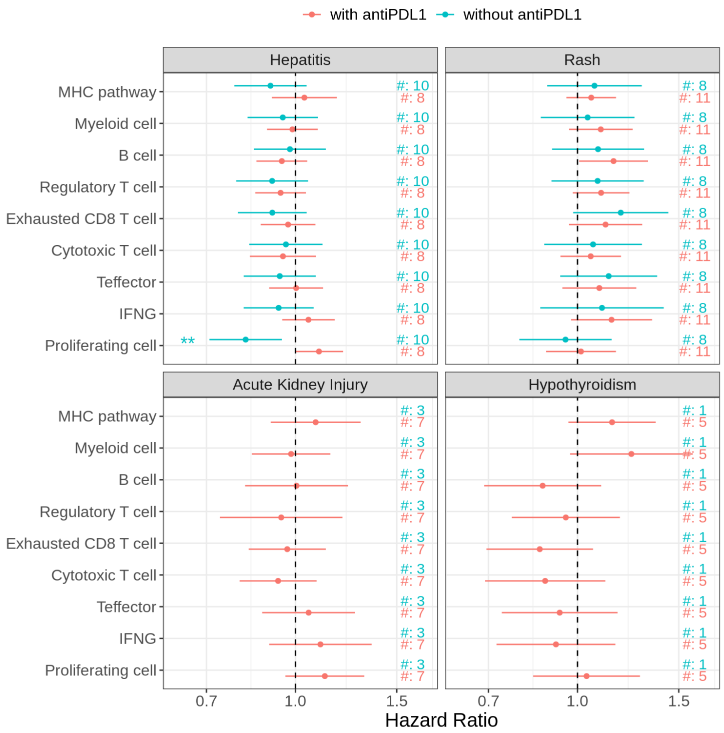


Figure 8: Association of TME gene signatures with irAEs stratified by Anti-PD-L1 combination status. Hazard ratios (HR) and 95% confidence intervals (CI) were obtained through a confounder-adjusted two-stage meta-analysis. #[number] indicates the number of studies included in the meta-analysis. ** indicates results with a p-value <0.01, and *** indicates results with a p-value <0.001.

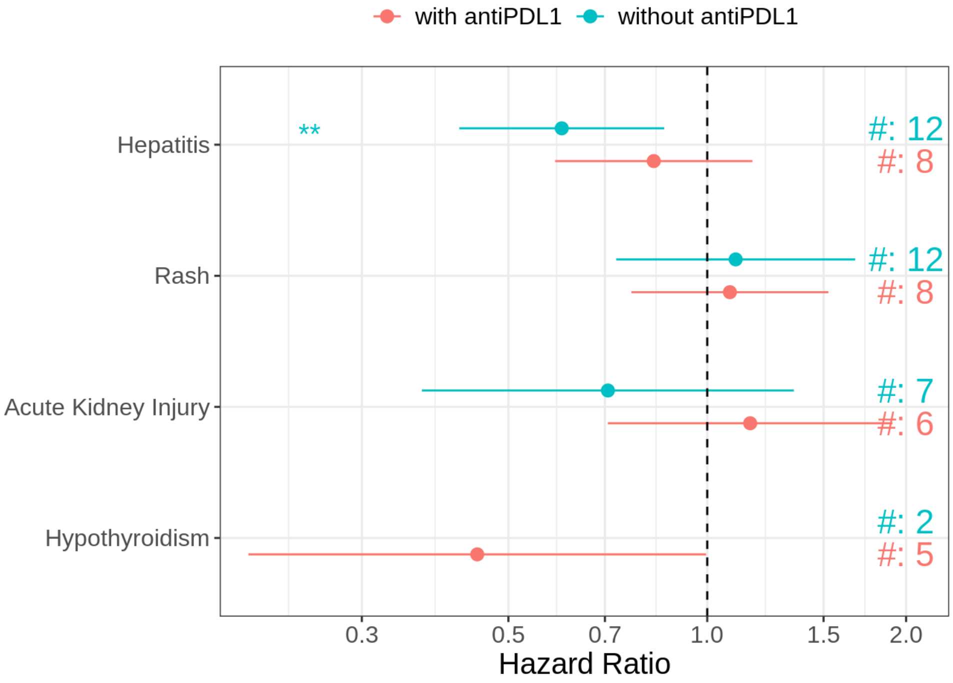


Figure 9: Association of previous CPI treatment with irAEs stratified by Anti-PD-L1 combination status. Hazard ratios (HR) and 95% confidence intervals (CI) were obtained through a confounder-adjusted two-stage meta-analysis. #[number] indicates the number of studies included in the meta-analysis. ** indicates results with a p-value <0.01, and *** indicates results with a p-value <0.001.

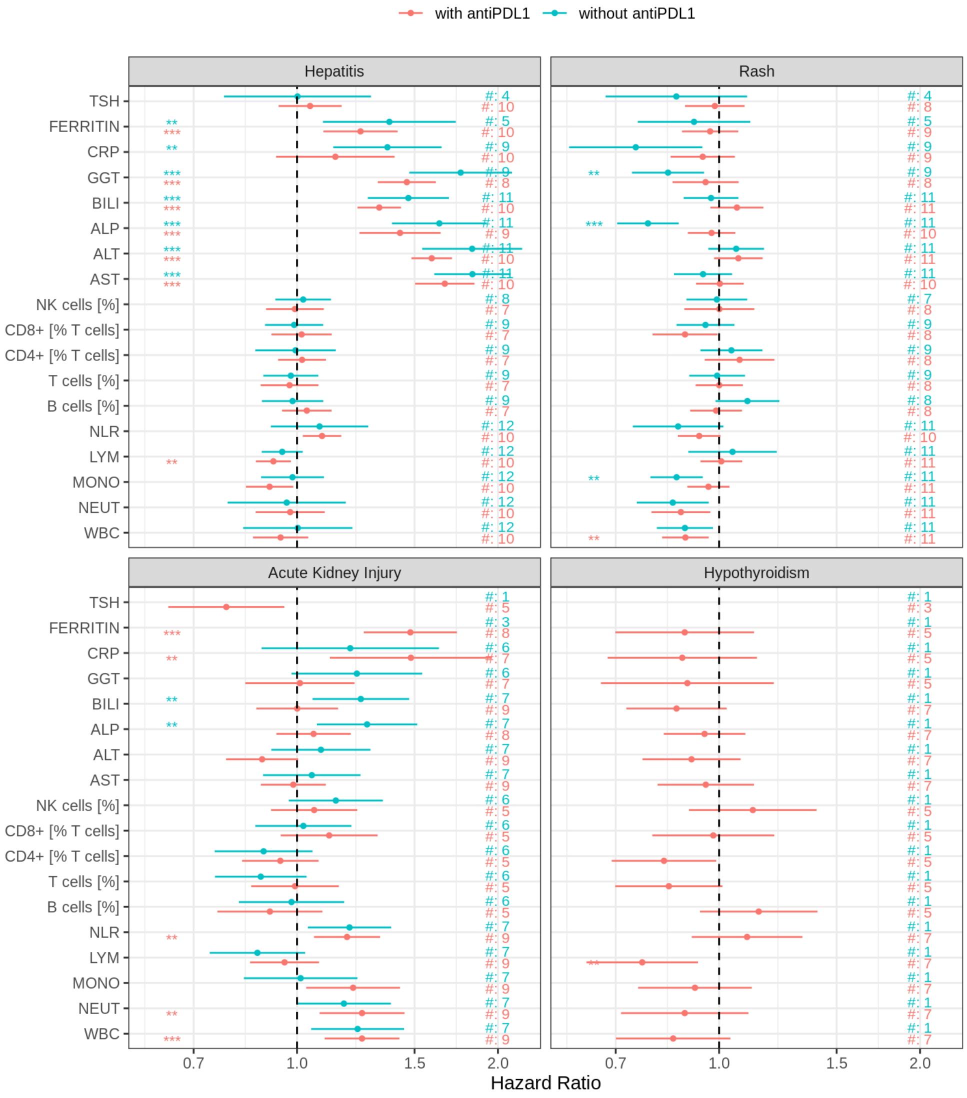


Figure 10: Association of soluble biomarkers with irAEs stratified by Anti-PD-L1 combination status. Hazard ratios (HR) and 95% confidence intervals (CI) were obtained through a confounder-adjusted two-stage meta-analysis. #[number] indicates the number of studies included in the meta-analysis. ** indicates results with a p-value <0.01, and *** indicates results with a p-value <0.001.

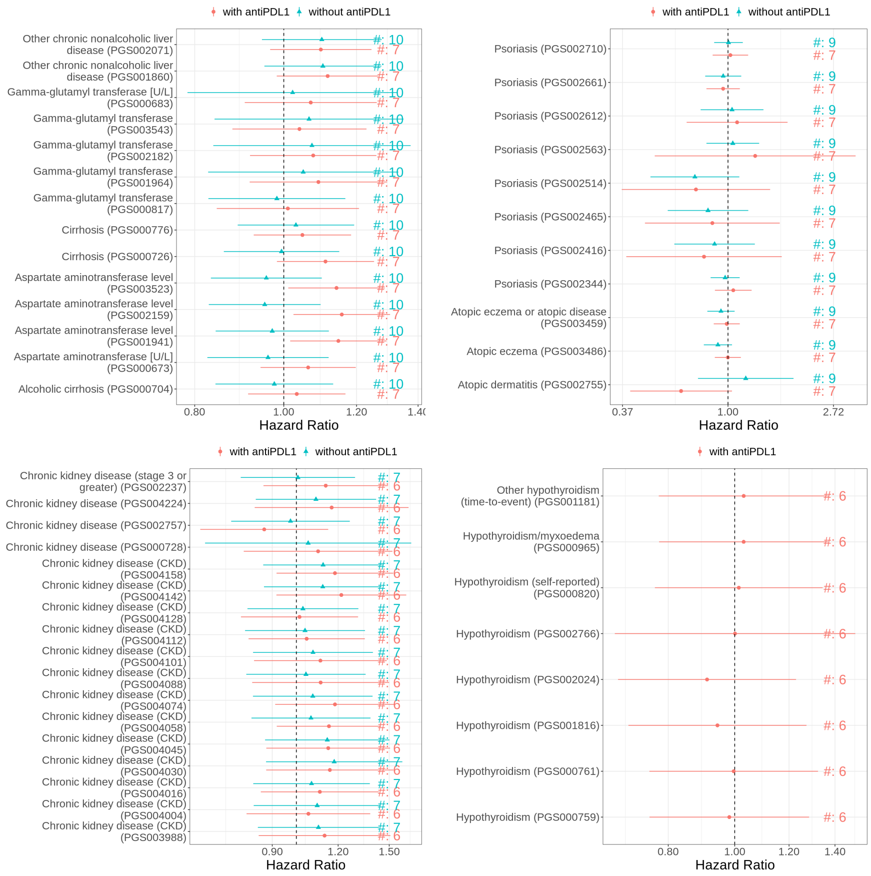


Figure 11: Association with PGS linked to irAEs stratified by Anti-PD-L1 combination status. Hazard ratios (HR) and 95% confidence intervals (CI) were obtained through a confounder-adjusted two-stage meta-analysis. #[number] indicates the number of studies included in the meta-analysis. ** indicates results with a p-value <0.01, and *** indicates results with a p-value <0.001.

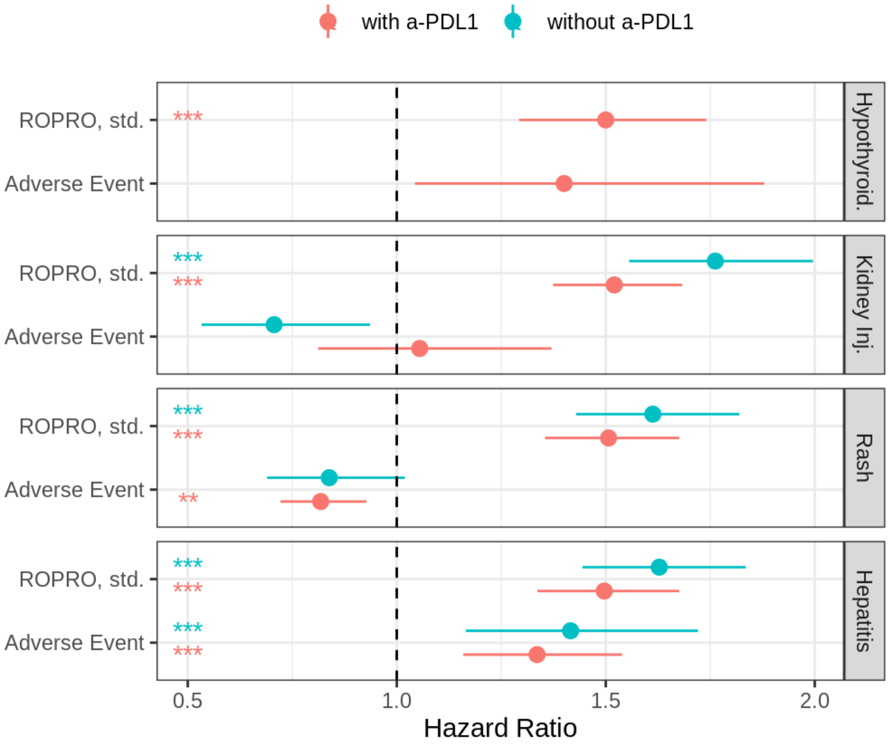


Figure 12: Association of irAEs and ROPRO with PFS stratified by Anti-PD-L1 combination status. Hazard ratios (HR) and 95% confidence intervals (CI) were obtained through bivariate two-stage meta-analysis treating PFS as the time-to-event endpoint and irAE and ROPRO as covariates. #[number] indicates the number of studies included in the meta-analysis. ** indicates results with a p-value <0.01, and *** indicates results with a p-value <0.001.

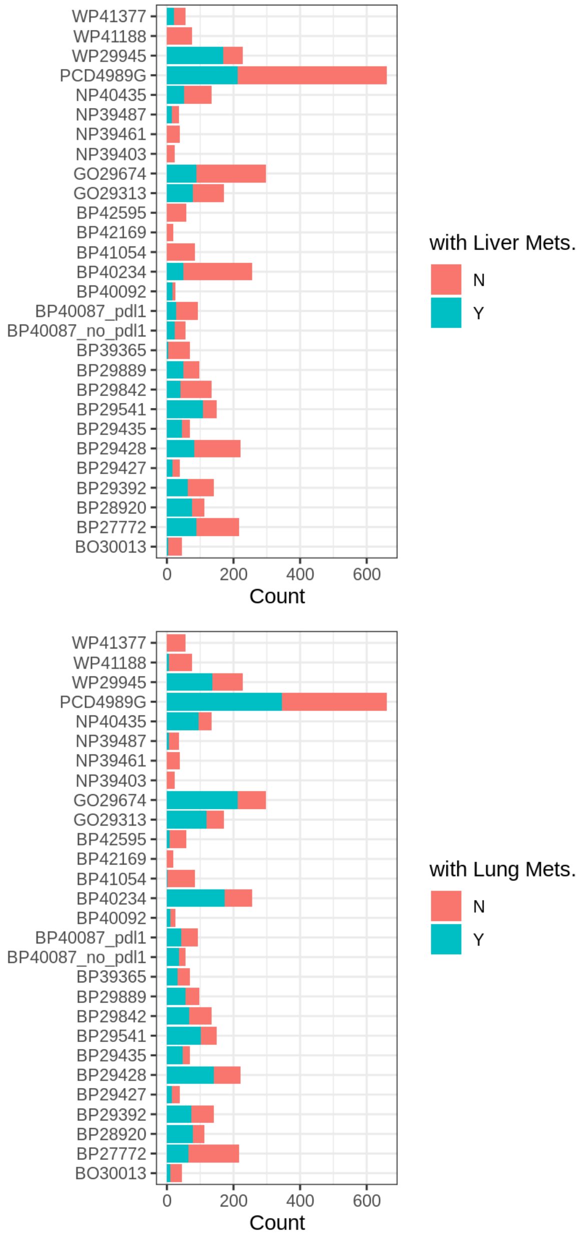


Figure 13: Distribution of tumor characteristics, including: the presence of liver and lung metastases (Liver/Lung Mets).

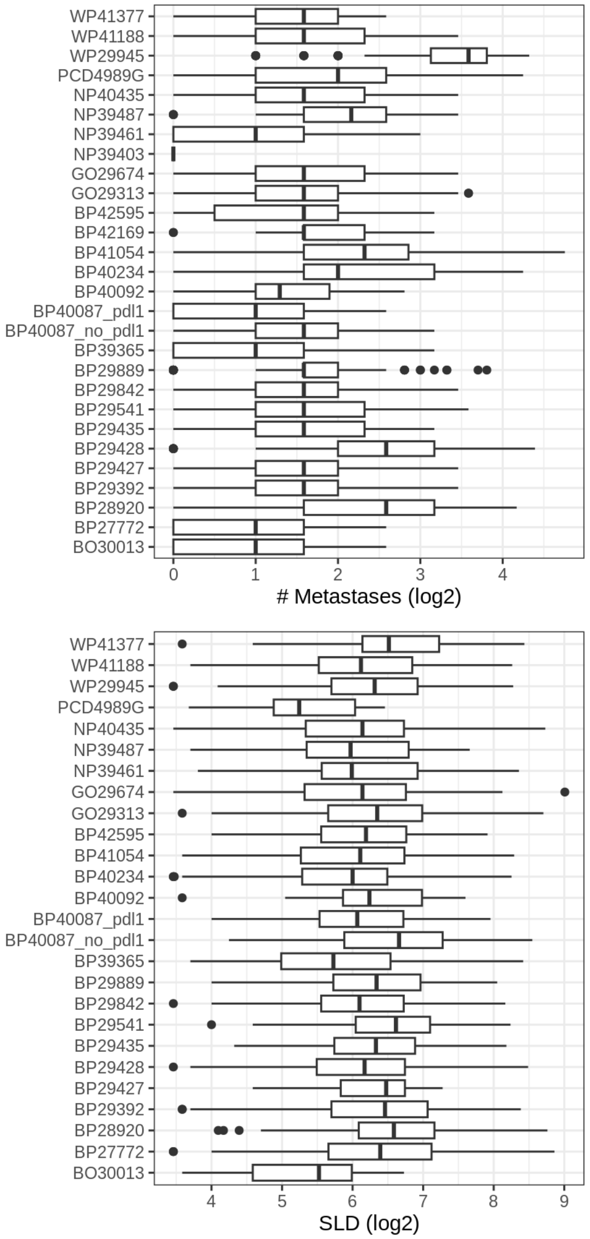


Figure 14: Distribution of tumor characteristics, including: the number of metastases (log2-transformed), and the sum of the largest diameters of target lesions (SLD, log2-transformed).

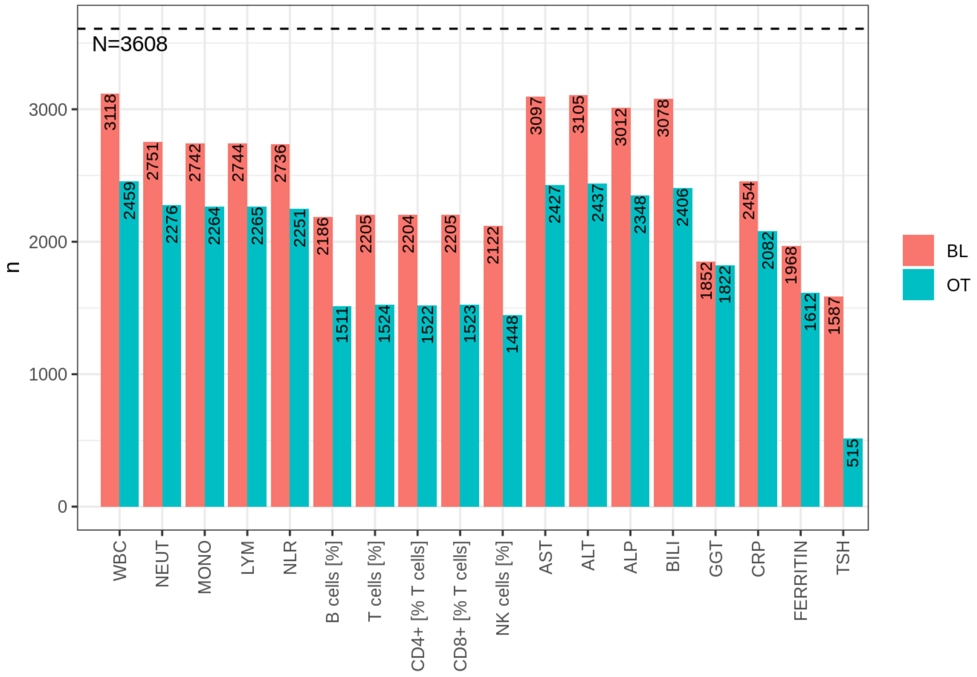


Figure 15: Sample size of soluble biomarkers: Bar plot depicting the number of patients with baseline samples (red) and the number of patients with at least one on-treatment sample (blue).

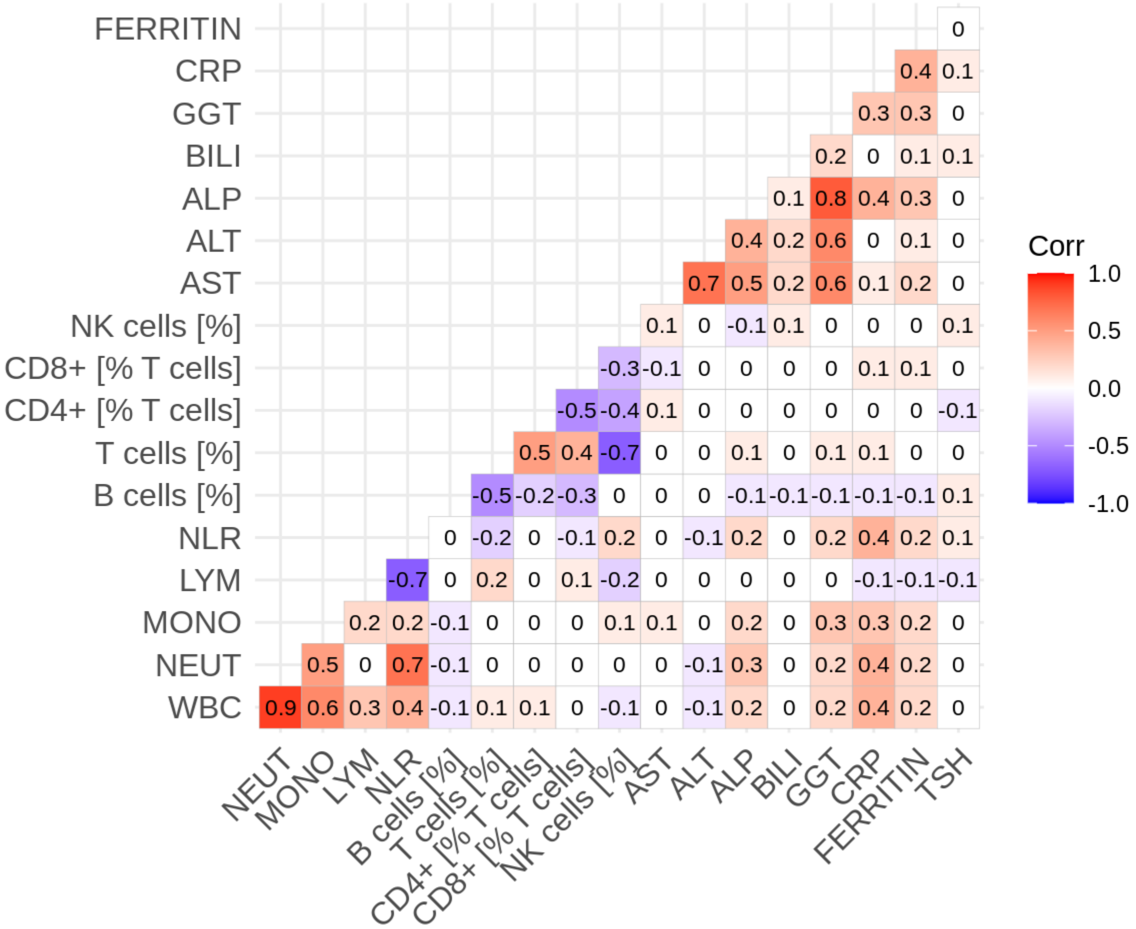


Figure 16: Correlation of soluble biomarkers: Correlation was calculated at the study level. The number and color indicate the median correlation across all studies.

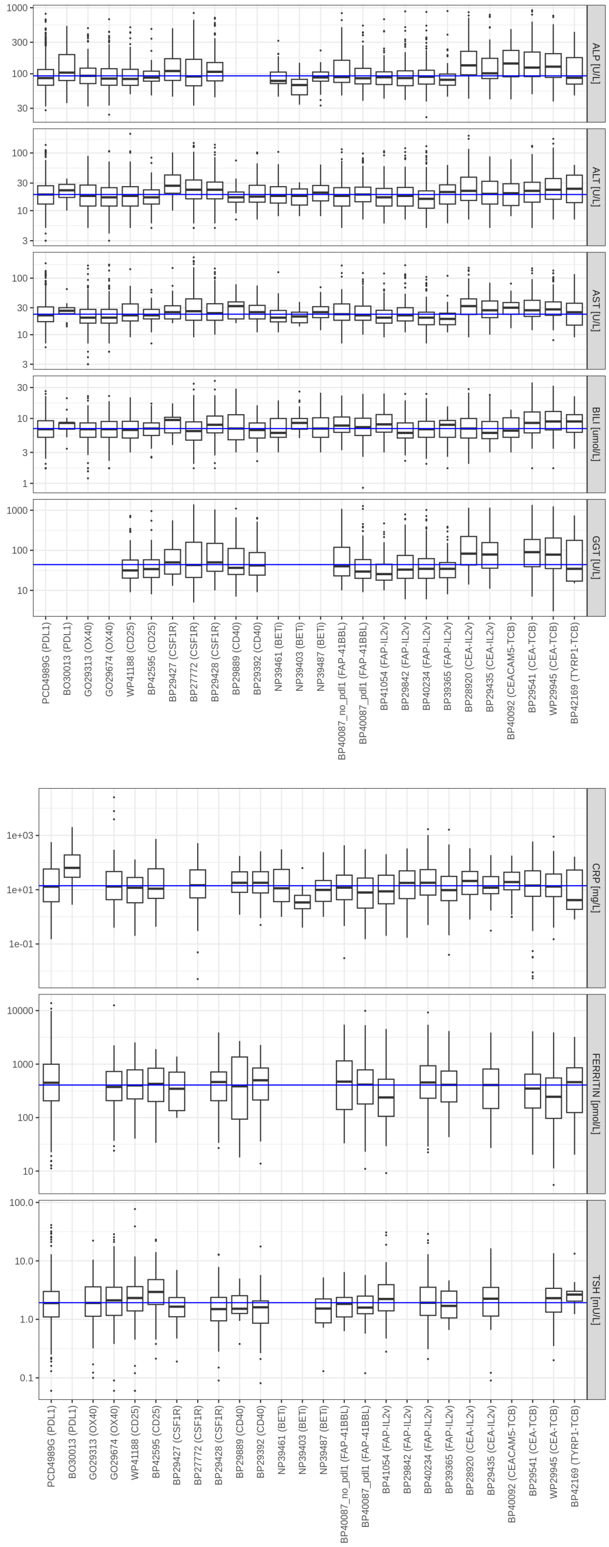


Figure 17: Distribution of soluble biomarkers: Boxplots illustrating the distribution of different biomarkers across studies. The blue line indicates the median across all studies.

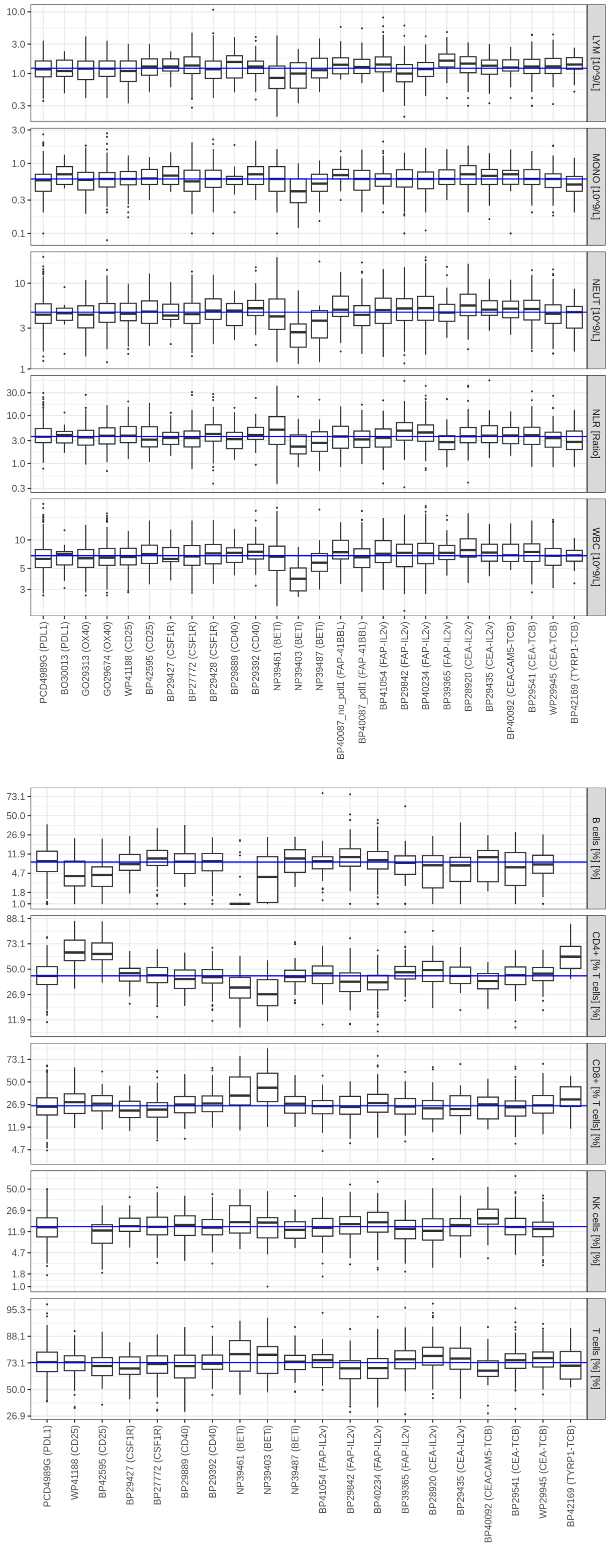


Figure 18: Distribution of soluble biomarkers: Boxplots illustrating the distribution of different biomarkers across studies. The blue line indicates the median across all studies.

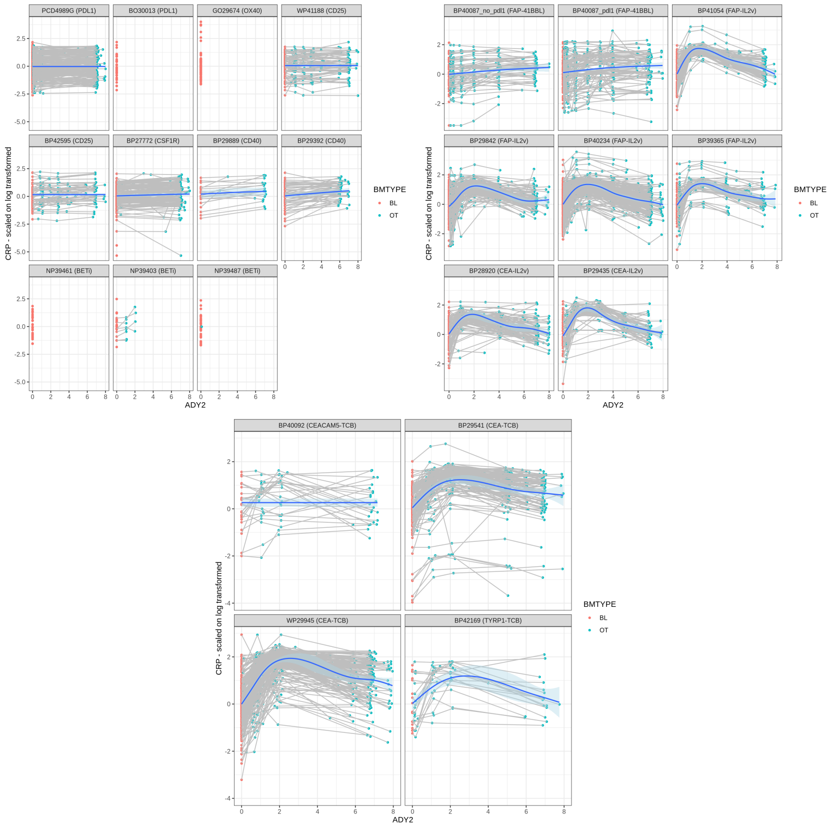


Figure 19: Figure depicting the longitudinal profile of CRP with blue dots representing on-treatment samples and red dots indicating baseline measurements. The blue line summarizes the study-level profile and is obtained using a smoothing technique.
